# Estimation of COVID-19 risk-stratified epidemiological parameters and policy implications for Los Angeles County through an integrated risk and stochastic epidemiological model

**DOI:** 10.1101/2020.12.11.20209627

**Authors:** Abigail L. Horn, Lai Jiang, Faith Washburn, Emil Hvitfeldt, Kayla de la Haye, William Nicholas, Paul Simon, Maryann Pentz, Wendy Cozen, Neeraj Sood, David V. Conti

**Author notes:** Corresponding author Email address (Abigail L. Horn).

## Abstract

**Summary:** *Background:* Health disparities have emerged with the COVID-19 epidemic because the risk of exposure to infection and the prevalence of risk factors for severe outcomes given infection vary within and between populations. However, estimated epidemic quantities such as rates of severe illness and death, the case fatality rate (CFR), and infection fatality rate (IFR), are often expressed in terms of aggregated population-level estimates due to the lack of epidemiological data at the refined subpopulation level. For public health policy makers to better address the pandemic, stratified estimates are necessary to investigate the potential outcomes of policy scenarios targeting specific subpopulations.

*Methods:* We develop a framework for using available data on the prevalence of COVID-19 risk factors (age, comorbidities, BMI, smoking status) in subpopulations, and epidemic dynamics at the population level and stratified by age, to estimate subpopulation-stratified probabilities of severe illness and the CFR (as deaths over observed infections) and IFR (as deaths over estimated total infections) across risk profiles representing all combinations of risk factors including age, comorbidities, obesity class, and smoking status. A dynamic epidemic model is integrated with a relative risk model to produce time-varying subpopulation-stratified estimates. The integrated model is used to analyze dynamic outcomes and parameters by population and subpopulation, and to simulate alternate policy scenarios that protect specific at-risk subpopulations or modify the population-wide transmission rate. The model is calibrated to data from the Los Angeles County population during the period March 1 - October 15 2020.

*Findings:* We estimate a rate of 0.23 (95% CI: 0.13,0.33) of infections observed before April 15, which increased over the epidemic course to 0.41 (0.11,0.69). Overall population-average IFR(*t*) estimates for LAC peaked at 0.77% (0.38%,1.15%) on May 15 and decreased to 0.55% (0.24%,0.90%) by October 15. The population-average IFR(*t*) stratified by age group varied extensively across subprofiles representing each combination of the additional risk factors considered (comorbidities, BMI, smoking). We found median IFRs ranging from 0.009%-0.04% in the youngest age group (0-19), from 0.1%-1.8% for those aged 20-44, 0.36%-4.3% for those aged 45-64, and 1.02%-5.42% for those aged 65+. In the group aged 65+ for which the rate of unobserved infections is likely much lower, we find median CFRs in the range 4.4%-23.45%. The initial societal lockdown period avoided overwhelming healthcare capacity and greatly reduced the observed death count. In comparative scenario analysis, alternative policies in which the population-wide transmission rate is reduced to a moderate and sustainable level of non-pharmaceutical interventions (NPIs) would not have been sufficient to avoid overwhelming healthcare capacity, and additionally would have exceeded the observed death count. Combining the moderate NPI policy with stringent protection of the at-risk subpopulation of individuals 65+ would have resulted in a death count similar to observed levels, but hospital counts would have approached capacity limits.

*Interpretation:* The risk of severe illness and death of COVID-19 varies tremendously across subpopulations and over time, suggesting that it is inappropriate to summarize epidemiological parameters for the entire population and epidemic time period. This includes variation not only across age groups, but also within age categories combined with other risk factors analyzed in this study (comorbidities, obesity status, smoking). In the policy analysis accounting for differences in IFR across risk groups in comparing the control of infections and protection of higher risk groups, we find that the strict initial lockdown period in LAC was effective because it both reduced overall transmission and protected individuals at greater risk, resulting in preventing both healthcare overload and deaths. While similar numbers of deaths as observed in LAC could have been achieved with a more moderate NPI policy combined with greater protection of individuals 65+, this would have come at the expense of overwhelming the healthcare system. In anticipation of a continued rise in cases in LAC this winter, policy makers need to consider the trade offs of various policy options on the numbers of the overall population that may become infected, severely ill, and that die when considering policies targeted at subpopulations at greatest risk of transmitting infection and at greatest risk for developing severe outcomes.

## Research in Context

### Evidence Before This Study

For public health policy makers to address the COVID-19 pandemic, stratified estimates are necessary to investigate the potential outcomes of policy scenarios targeting specific subpopulations. However, estimated epidemic quantities such as rates of severe illness and death, the case fatality rate (CFR) and the infection fatality rate (IFR) have been expressed in terms of aggregated population-level estimates or estimates by age groups alone, due to the lack of epidemiological data at the refined subpopulation level. We searched PubMed for articles published in English from inception to November 25, 2020 with the keywords “covid-19” AND “ifr” OR “infection fatality risk” OR “infection fatality rate” AND “age” AND “obesity” OR “comorbidity” OR “smoking”, which identified 31 results. We found a few estimates of odds ratios and hazard ratios for each potential risk factor controlling for other risk factors, but found no estimates of case fatality rate or infection fatality rate by age and another moderator.

### Added Value of This Study

Conventionally, estimates of risk effects and outcomes given combinations of conditions for refined subpopulations are obtained through access to individual-level data and the application of multiple regression techniques. At the time of this study, individual-level COVID-19 data were not widely available nor sampled in an appropriate manner to avoid substantial bias. In the absence of access to appropriate individual-level data, we develop a framework for using available data on the prevalence of COVID-19 risk factors (age, comorbidities, BMI, smoking status) in subpopulations, and epidemic dynamics at the population-level and stratified by age, to estimate subpopulation-stratified probabilities of severe illness and the CFR and IFR (accounting for estimated total infections) across multiple combinations of risk factors. A dynamic epidemic model is integrated with a relative risk modeling approach to produce time-varying subpopulation-stratified estimates. The integrated model allows analyzing dynamic outcomes and parameters by population and subpopulation, and simulating alternate policy scenarios that protect specific at-risk subpopulations and modify the population-wide transmission rate. The model is calibrated to data from Los Angeles County from the period March 1 - October 15, 2020.

### Implications of All the Available Evidence

Results highlight the value of strata-specific estimates in anticipating disparities in the impact of the epidemic and the efficacy of targeted subpopulation-level policy interventions. The risk of severe illness given infection and the CFR and IFR vary tremendously across subgroups within a population and have changed over time, suggesting that it may be inappropriate to reduce the variation in risk to static, single summary measures for the entire population. This includes variation not only across age groups, but also within age categories combined with other risk factors analyzed in this study (comorbidities, obesity status, smoking). The fact that the risk of severe illness and the CFR and IFR vary tremendously across subpopulations raises the question of whether protecting certain subpopulations would have achieved similar results in infections, hospitalizations, and deaths as policies applied population-wide to reduce transmission, including the initial lockdown period from March 19 to May 8. We investigated this question through counter-factual simulation, comparing observed trends with alternate policy scenarios modifying the population-wide transmission rate or protecting specific at-risk subpopulations. We find the strict initial lockdown period in LAC was effective because it both reduced overall transmission and protected individuals at greater risk, preventing both healthcare overload and deaths. While similar numbers of deaths as observed in LAC could have been achieved while avoiding initial societal lockdown through the implementation of a more moderate policy with regard to transmission reduction through non-pharmaceutical interventions (NPIs) combined with greater protection of individuals 65+, this would have come at the potential expense of overwhelming the healthcare system. These results illustrate the value in policy analysis of a reproducible model-based method for producing subpopulation-level disease severity and IFR estimates without access to individual data, which can be applied in other locations and policy settings such as the evaluation of vaccination policy focused on targeting at-risk sub-groups.

## 1. Introduction

Health disparities have emerged with the COVID-19 epidemic because the risk of exposure to infection and the prevalence of risk factors for severe outcomes given infection vary within and between populations and over time [1, 2, 3, 4, 5, 6, 7]. For public health policy makers to better address the pandemic, models reporting stratified estimates are necessary to investigate the potential outcomes of policy scenarios targeting specific subpopulations. However, estimated epidemic quantities such as rates of severe illness and death, the case fatality rate (*CFR*), and the infection fatality rate (*IFR*) are often expressed in terms of aggregated population-level estimates or by age group alone due to the lack of epidemiological data at the refined subpopulation level [8, 9, 10]. While data may be available for single risk factor strata such as by age [11], data on subpopulations representing individuals with combinations of risk factors are not reported or available. Conventionally, estimates of risk effects and outcomes given combinations of conditions are obtained through access to individual-level data and the application of multiple regression techniques [12, 5]. At the time of this study, individual-level COVID-19 data were not widely available nor sampled in an appropriate manner to avoid substantial bias [13].

In this paper we develop a model that produces stratified estimates of illness severity and death for subpopulations representing individuals with combinations of risk factors from available dynamic epidemiological data at the aggregated population level [11]. In the absence of access to individual-level data, we apply a statistical technique developed for conditional analysis of marginal summary statistics [14] to obtain estimates of the conditional effects of combinations of risk factors for COVID-19 on the probability of severe illness and death using data from published studies reporting the marginal effects of individual risk factors [3, 2]. We consider the risk factors age, existing comorbidities, obesity, and smoking. Separately, we develop a stochastic epidemic model that estimates time-varying probabilities of hospitalization, ICU admission, and death given infection at the population level. We integrate the conditional risk effects and the population-level probabilities, together with available dynamic data on the prevalence of infections stratified by age, in a framework that estimates the probability of advancing to each stage of disease, CFRs, and IFRs, stratified across all plausible combinations of the modeled risk factors.

The integrated model allows the analysis of dynamic outcomes and parameters by population and subpopulation. Focusing on Los Angeles County (LAC), the most populous and one of the most diverse counties in the United States, we analyze the estimated overall and risk-stratified time-varying illness severity probabilities, CFRs, and IFRs in relation to the epidemic timecourse and implemented policy decisions. We focus on the time period March 1 through October 15, marking the end of what we characterize as a second wave of the epidemic and before a third wave of rising infection began. Through simulation, we compare observed trends with alternate policy scenarios protecting specific at-risk subpopulations or modifying the level of non-pharmaceutical intervention (NPIs) implementation. Results highlight the value of strata-specific estimates in understanding disparities in the impact of the epidemic, and the efficacy of targeted subpopulation-level policy interventions in LAC.

## 2. Methods

We first developed a single-population stochastic dynamic epidemic model that accounts for observed and unobserved transmission of COVID-19 and trajectories through the healthcare system with hospitalization, ICU admission, and death. Using Bayesian methods for parameter estimation and uncertainty quantification, we estimated the population-average time-varying probabilities of transitions between the infected, hospitalized, ICU, mechanical ventilation, death, and recovery compartments, and the resulting population-average time-varying case fatality rate (*CFR*, defined as deaths over observed infections) and infection fatality rate (*IFR*, defined as deaths over all infections) (Section 2.1). In parallel, we used available data from published studies on the marginal effects of individual risk factors (age, existing comorbidities, obesity, smoking) to calculate conditional risk effects estimates for three models: (1) hospitalization given infection, (2) ICU admission given hospitalization, and (3) death given hospitalization. The conditional risk estimates were integrated with the corresponding probability estimates from the dynamic epidemic model to create a *risk model* (Section 2.2). The *risk model* enables us to estimate, stratified across 39 combinations of modeled risk factors (i.e. *risk profiles*), the probability of each stage of disease given infection within LAC. Furthermore, we integrate the time-varying stratified probability of each stage of disease with the time-series of observed infections, estimated total infections including observed and unobserved, deaths, together with available data on the prevalence of infections stratified by age, to estimate the risk profile-stratified CFR and IFR across time.

### 2.1. Epidemic model

We develop a model of COVID-19 t.ransmission in a single, homogeneously-mixed population divided into nine compartments representing different disease states (Figure 1). Compartments relating to the transmission of infection are the widely-used susceptible, exposed (latent but not yet infectious), infectious and observed (*I*), and recovered classes. We also include a compartment representing unobserved and/or unconfirmed infections (*A*). We model healthcare utilization and outcome at a more granular level by including compartments representing individuals that are in hospital (*H*), in ICU care (*Q*), undergoing mechanical ventilation support (*V*), and that die (*D*). Each individual can only be in one state at each point in time with the exception of the mechanical ventilation support class, which is viewed as a proxy with error for the in-ICU class. We assume that new infections are created only by individuals in the infected classes (*I* and *A*), and that individuals in all other compartments, including in hospital, do not contribute to transmission.

**Figure 1:**
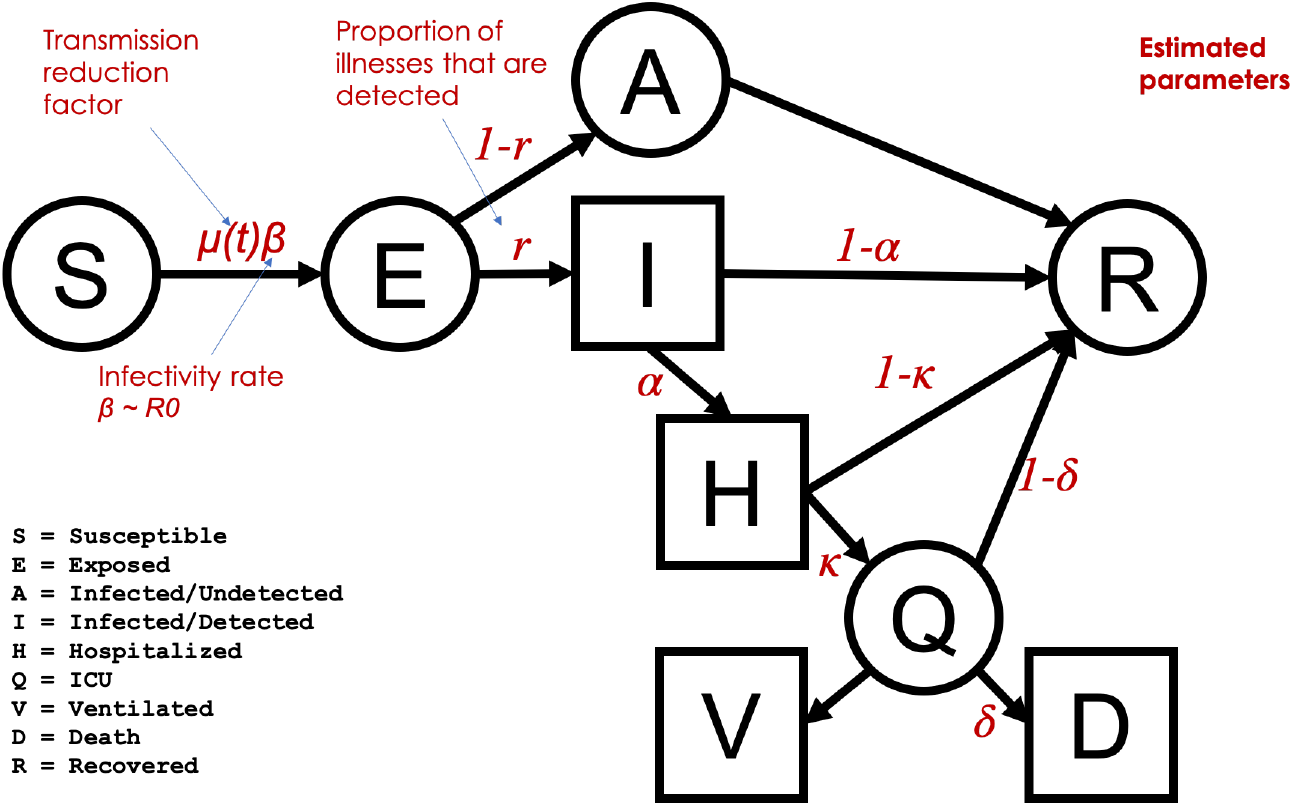
Epidemic model structure and key estimated parameters. Model compartments with available data are represented as square compartments

To model this dynamical state system we employ a discrete-time approximation to the corresponding stochastic continuous-time Markov process in which transitions of individuals between disease stages are seen as stochastic movements between the corresponding population compartments with random transition rates [15, 16]. This model keeps track of the number of individuals in each compartment and the flows of individuals transitioning between compartments through a set of coupled discrete-time multinomial counting processes with transmission rates defined by Poisson processes. To simulate from this system we employ a Euler numerical scheme for Markov process models [15]. For more details see appendix pp 1-6.

The basic reproductive number, *R*0, defined as the mean number of secondary cases generated by a typical infectious individual on each day in a full susceptible population [17], is a function of model parameters including the transmission rate *β*(*t*) [18] (appendix pp 7-8), defined as the average number of individuals that an infected individual will infect per day. We introduce an additional time-varying parameter, *µ*(*t*), representing modification to the original *β*_0_ and equivalently *R*0, which we call the *transmission reduction factor*. The transmission reduction factor allows us to estimate changes in the transmission rate over time and to explicitly model changes in the transmission rate to simulate different policy scenarios. We define the time-varying effective reproductive number as *R*(*t*) = *µ*(*t*)*R*0, i.e. the initial value of the basic reproductive number at the beginning of the pandemic period before interventions multiplied by the time-varying transmission reduction factor (appendix p 6).

#### 2.1.1. Parameter Estimation

All transition rate parameters (e.g., the time between exposure and infectiousness) are modeled as fixed values taken directly from published literature (appendix p 3). The model has six unknown parameters, *θ* = {*β*(*t*), *r*(*t*), *α*_*t*_, *κ*_*t*_, *δ*_*t*_, *p*_*V*_}, which we estimate using Approximate Bayesian Computation (ABC) techniques using multiple data sources to specify informative prior parameter distributions. The prior distribution for *R*0 is informed by values estimated from other published studies on COVID-19 [19, 20]. We use geolocation trace data from smartphones, i.e. mobility data, to inform both the magnitude and the timing of inflection points in the transmission reduction factor *µ*(*t*). We incorporate data for LAC provided by Unacast [21] on reductions in distances travelled and encounter rates [22]. The prior distribution for the fraction of observed cases out of all infections, *r*(*t*), was informed by results of a CDC study reporting seroprevalence surveys across 10 communities in March - early May for dates within that time period, and was allowed to vary more widely for dates beyond that time period [23] (appendix pp 8-11).

Changes in the probabilities of each disease stage over time are determined by the changing risk profile of the infected population. The only available infection data for LAC by risk factor was for age [11]. We therefore used the distribution of infections by age to inform the timescale and shape of the linear function modeling *α*_*t*_, *κ*_*t*_, and *δ*_*t*_ over time. Prior distributions for each probability on key dates were modeled as normal distributions with means informed by the ratios of the observed numbers of infections, hospitalizations, and deaths in LAC (appendix p 11).

The model was fit to the daily and cumulative count of infections (observed), hospitalizations, individuals undergoing ventilation support, and deaths in LAC from a data set from the LACDPH, updated daily by the COVID-19 Outbreak Data Coordination Team [11](appendix p 12). Using ABC on multiple parameters simultaneously produces joint posterior estimates over all parameters. The CFR(*t*) and IFR(*t*) were calculated as estimated deaths over estimated cumulative observed infections or estimated cumulative total infections, respectively. To provide uncertainty estimates, we simulate trajectories with parameter values coming from the joint posterior distribution and aggregate simulations from 1000 jointly estimated parameter sets. For each parameter set, we aggregate simulations for 100 stochastic epidemic model realizations. We pool together all simulations and report their median and 95% credible intervals (CI) as the 2.5th/97.5th quantiles of realizations. This procedure quantifies uncertainty from two sources: variability due to joint estimated parameter values, and variability due to the stochastic variability between model runs with the same parameter values.

### 2.2. Risk model

Using studies reporting the risk of severe COVID-19 outcomes given individual risk factors [3, 2], we construct a logistic regression model to estimate the probability of infection for individuals with combinations of risk factors. Specifically, we estimate three models for: (1) the probability that an individual is admitted to hospital given acquired (observed) infection *P*_*t*_(*H*|*I*), (2) the probability the individual is admitted to the ICU given admittance to hospital *P*_*t*_(*Q*|*H*), and (3) the probability that the individual dies given being admitted to the ICU *P*_*t*_(*D*|*Q*). These probabilities correspond to the epidemic model estimated parameters *α*_*t*_, *κ*_*t*_, and *δ*_*t*_, respectively. Each of these regression models includes indicator variables for the presence or absence of specific risk factors.

The risk factors included in our analysis are age, body mass index (BMI), smoking status, and any comorbidity. The comorbidities included are diabetes, hypertension, chronic obstructive pulmonary disease (COPD), hepatitis B, coronary heart disease, stroke, cancer and chronic kidney disease. We modeled age and BMI as an ordinal variable and assume an additive effect of both age and BMI on the three outcomes. Age was categorized within four groups: 0 *−* 19, 20 *−* 44, 45 *−* 64, and 65+, and BMI was categorized in three groups according to obesity classes: Class 1 (no obesity) 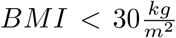; Class 2 (obesity), 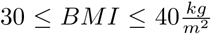; Class 3 (severe obesity), 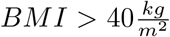. Any comorbidity and smoking status were modeled as binary variables.

We estimate the conditional risk effects corresponding to each factor for each risk model using marginal effects estimates available from reported studies and a method called the joint analysis of marginal summary statistics (JAM) [14]. JAM uses two pieces of information: (i) the marginal effect estimates between risk factors and the outcome and (ii) a reference correlation structure between the risk factors. For information informing (i) we obtain the marginal log RR between individual risk factors and COVID-19 illness severity from peer-reviewed published COVID-19 studies [3, 2] (left column of Table 2). For (ii), we use data from The National Health and Nutrition Examination Survey (NHANES) from 2017-2018 [24] (details in appendix pp 13-14).

To construct a model to estimate *P*_*t*_(*H*|*I*), *P*_*t*_(*Q*|*H*), and *P*_*t*_(*D*|*Q*), we combine in a logistic model the 39 risk profiles representing all plausible linear combinations of the risk factors specified in a mean centered design matrix, **X**; and their corresponding conditional log RR obtained from JAM, 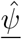; with specified intercepts set to the estimated probabilities from the epidemic model (Section 2.1) for 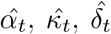, respectively (appendix pp 16-17). For example, to estimate the vector of probabilities of hospitalization given infection for all risk profiles we use 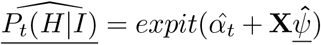. The reference profile are individuals age 0 *−* 19 with no comorbidity, 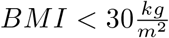, and non-smoking. The specification of the frequency of each risk profile at each stage of disease for the mean centered design matrix is obtained from available LAC data on the prevalence of each age group in infections [11] and an estimate of the frequency of each risk profile within the overall population based on prevalence data for individual risk factors [25, 26], updated at each stage of disease (appendix pp 14-16). We report estimates at three time points: May 15, August 1, and October 15, 2020.

### 2.3. Calculating Risk Profile Stratified CFR(t) and IFR(t)

To calculate the time-varying CFR(*t*) and IFR(*t*) for each risk profile, the estimated frequency of each profile in the infected population and in the deceased population (obtained from the risk model) are multiplied by each value of the estimated cumulative number of observed infections (*I*) or total infections (*A*), and deaths (*D*), respectively. We find the CFR(*t*) and IFR(*t*) for each model realization as the number of deaths over observed infections, and number of deaths over total infections, respectively. Repeating across the 1000 model realizations achieves the 95% CI (appendix pp 18-19).

### 2.4. Scenario Analysis

We implement scenarios modeling the protection of at-risk populations and changes to the population-wide transmission rate at different times. We increase or decrease the value of the reproductive number *R*(*t*) to reflect different levels of non-pharmaceutical interventions (NPIs), which could include measures such as physical distancing and/or mask adherence. Specifically, we model three levels of population-wide transmission rates: (1) *NPIs=Observed* implements the observed (epidemic model-estimated) *R*(*t*) throughout the epidemic in LAC.(2) *NPIs=Moderate* implements an *R*(*t*) equal to the estimated maximum value reached in LAC after the initial lockdown restrictions were eased (Section 3.1). Although this maximum value for *R*(*t*) was reached on May 15, in this scenario we decrease *R*(*t*) from the initial *R*0 to this maximum *R*(*t*) value between March 12 and March 27 to reflect no community lockdown implemented. (3) *NPIs=None* implements *R*(*t*) = *R*0 throughout the time interval March - October 2020, representing a baseline scenario in which no actions or behaviors reduce the native *R*0. The trend of *R*(*t*) modeled for the three levels is shown in Figure 4a.

To simulate scenarios protecting at-risk populations we focus on the shielding of individuals aged 65+ and calculate values of the probabilities of each stage of disease given infection (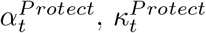, and 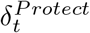) over time with the subpopulation of individuals 65+ removed. We isolate a fraction of individuals 65+ above the observed prevalence of this subpopulation in the infected population. We model three levels: (1) *Protect=Observed* models the trend in the probabilities of severe illness estimated by the epidemic model. We call this the “*Protect=Observed* “rather then “*Protect=None*” level because it follows the natural behaviorally-adapted reduction of individuals 65+ in the infected population (see Section **??**). (2) *Protect=High* isolates all individuals 65+, approximately 11% of the LAC population, from infection and subsequent stages of disease. We implement this scenario by calculating the population-average 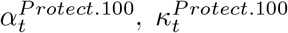 and 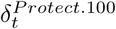 with individuals 65+ removed along the same timeline as the observed trend in *α*_*t*_, *κ*_*t*_, and *δ*_*t*_. The frequency of each risk profile in infections is made to reflect the age distribution over infections as observed in LAC data (following the approach described in Section 2.2), renormalized after profiles including individuals 65+ are removed. With the renormalized frequency distribution of each profile over infections, we use the risk model (Section 2.2) to find the population-average adjusted values. The timeline follows that for the observed trend in *α*_*t*_, *κ*_*t*_, and *δ*_*t*_. (3) *Protect=Moderate* isolates 50% of individuals 65+ from community transmission and subsequent stages of disease from the observed distribution of each age group over infections on each date and calculates the population-average adjusted 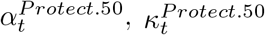, and 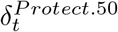, following the approach for the *Protect=High* scenario. The trend of the probabilities of each stage of disease given infection modeled for the three levels is shown in Figure 4b (full details in appendix pp 19-21).

We implement nine scenarios representing all combinations of the three *NPI* and three *Protect* settings. For each scenario, we simulate the model with the estimated parameter values for *R*0, *r*(*t*), *p*_*V*_, and starting time *t*_0_ with the remaining parameters implemented as described above.

## 3. Results

### 3.1. Model estimates and epidemic trends in LAC

#### Model fits

Figure 2 summarizes the epidemic model fit with COVID-19 data for LAC from March 1 through October 15 2020 for all disease states across multiple views: New cases, representing new daily incidence; the current number in a compartment at a specific date, relevant for understanding current prevalence rates and comparing with healthcare capacity limitations; and cumulative counts until a specific date. Observed data for available compartments are plotted as black dots. The figure demonstrates that good model fits are achieved in all compartments across time.

**Figure 2:**
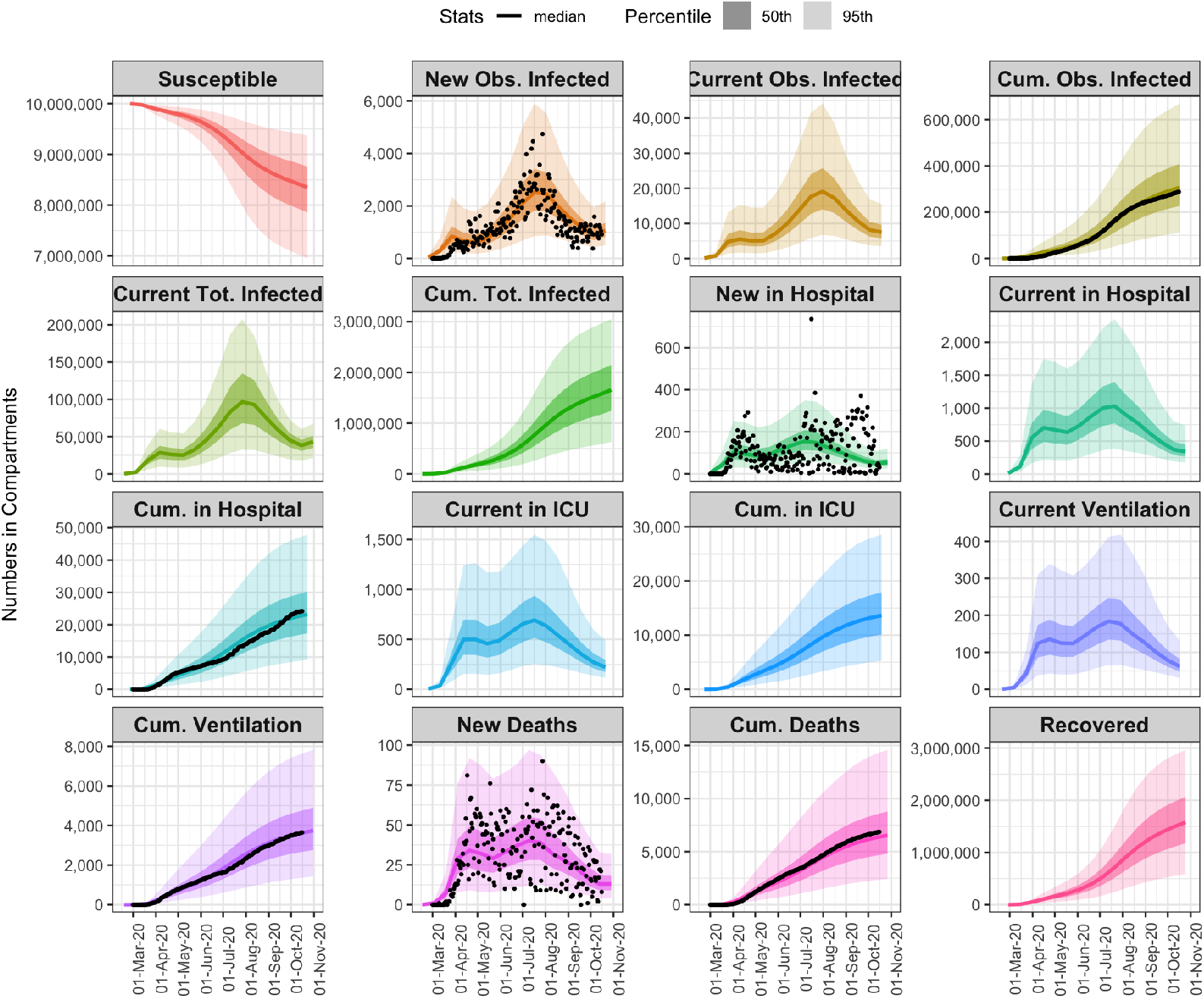
Summary of the epidemic model fit with COVID-19 data for Los Angeles, for all state variables, across multiple views: New cases, representing new daily incidence, current number in a compartment at a specific date, and cumulative counts. Available observed data (for new and cumulative counts)are plotted as black dots. Estimates are shown as the median number in compartments over time, with the 50% (darker) and 95% (lighter) CI.

#### Epidemic timecourse in LAC

The LA City Mayor’s Office distinguishes between three stages of the COVID-19 epidemic in LA City and County relating to policy response measures implemented following the orders of the County Health Officer: Stage I, March 19 - May 7: the initial shutdown; Stage II, May 8 - June 11: the first steps towards reopening; Stage III, June 12 - October 15 (and beyond): greater reopening followed by “modifications” closing higher risk settings (including bars and indoor seating in restaurants) [27, 28]. The start of the school year on August 18, although virtual, marked a change in activity level and is also depicted. We characterize three waves of the epidemic occurring across these stages: a first wave, March 1 - May 6, occurring between Stage I and the beginning of Stage II and peaking on April 1; a second and much larger wave, May 7 - October 14, beginning with Stage II and peaking during Stage III on July 30; and a third wave that began on October 15 and which is outside the scope of this paper. Figures 3a-3f characterize the epidemic course and estimated model parameters relative to these policy stages and epidemic waves. A full time course of the epidemic and policy decisions in LAC can be found at [29].

**Figure 3:**
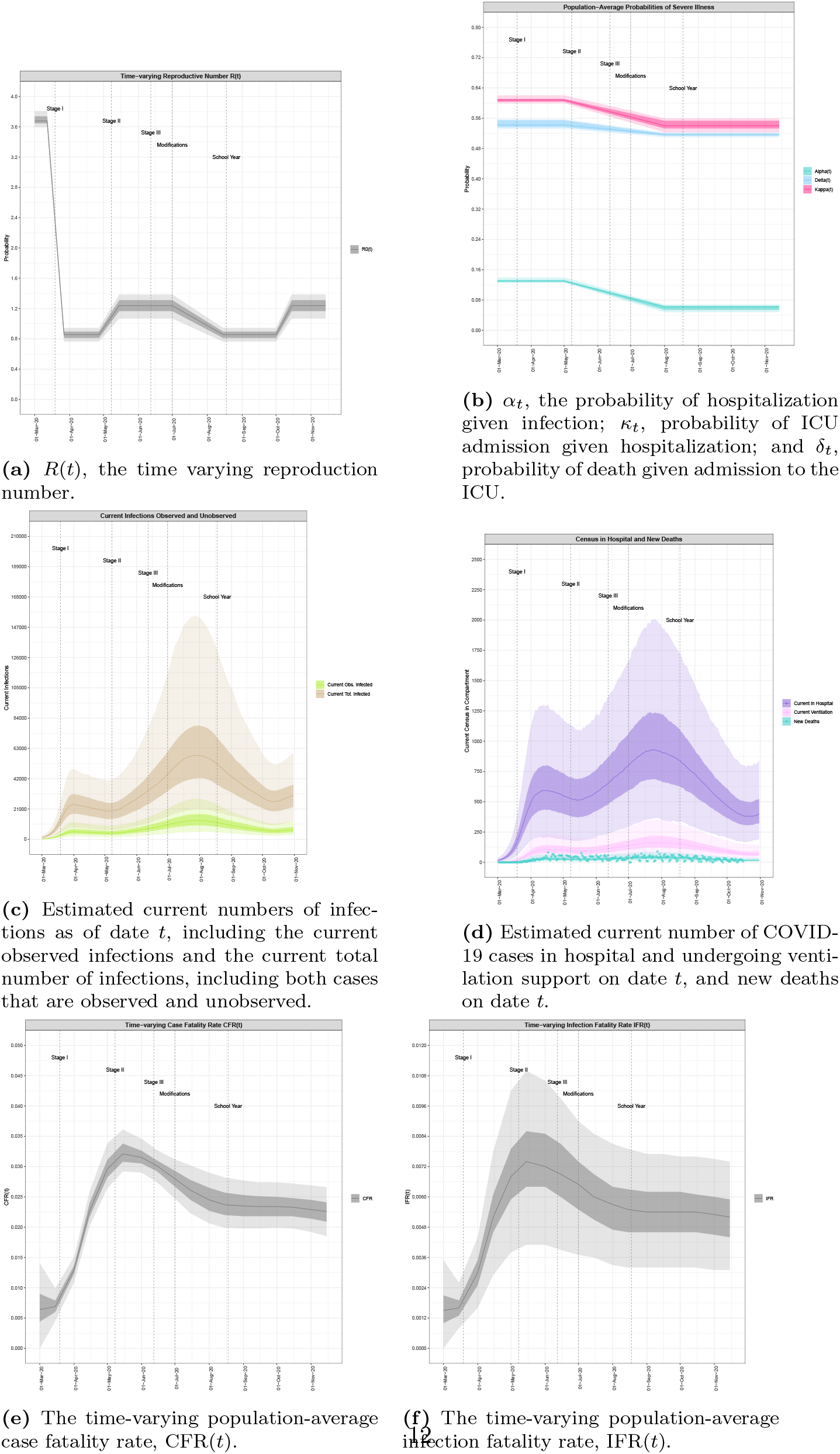
Timeseries of model-estimated parameters and compartmental variables relative to COVID-19 policy decisions in LAC. Model-estimated median curves are plotted along with the 50th% (dark shading) and 95% CI (light shading). Data is not available for current numbers but is plotted for the variable available (new deaths).

We estimate that for most of Stage I, the overall observation rate was *r*(*t*) = 0.23 (95% CI: (0.13,0.33) of all infections observed. Beginning in mid-April, the observation rate began to steadily increase until levelling off at a value of *r*(*t*) = 0.41 (0.11,0.69) by August 15. In the initial period of the outbreak before public behavior began to change and policy interventions were implemented, we estimate the basic reproduction number was *R*0=3.675 (3.627,3.729). During the period from March 12 to March 27, beginning just before Stage I was implemented, we estimate a reduction to an *R*(*t*) of 0.859 (0.758,0.947). The corresponding reduction in transmission led to a levelling off at 30 127 (39 039, 58 842) estimated current total infections on April 1 and subsequent decrease until May 6, followed by hospitalizations, ICU admissions, and deaths. Notably, *R*(*t*) began to rise again following increasing mobility behavior on April 27, almost two weeks before the Stage II reopening policy was implemented, portending the increase in infections to follow.

Throughout Stage II and the beginning of Stage III, we estimate an increasing trend in *R*(*t*), reaching 1.264 (1.052,1.378) around the time that the “modifications” (re-closures) were implemented at the beginning of July. The increasing *R*(*t*) during this period was followed by a second exponential growth phase in infections, hospitalizations, ICU admissions, and deaths. *R*(*t*) began to decrease following the implementation of the “modifications,” and by the start of the school year in mid-August (virtually) had returned to the Stage I value where it plateaued until a new rise at the beginning of October. Estimated current total infections continued to increase until a peak of 93 720 (34 404, 184 084) on July 30, reflecting the expected delay between reductions in *R*(*t*) and the infection rate. Current total infections followed a decreasing trend until October 15, marking the beginning the third wave of the epidemic. Hospitalizations and ICU admissions followed the infection rate and reached much larger second peaks in late July before decreasing again until reaching new minimums in mid-October. Importantly, even at upper 95%CI of the peak, the current census of cases in hospital, ICU, and undergoing ventilation support remain well below the capacity limits of approximately 4 000 hospital beds and 2 245 ICU beds in LAC [11].

Although current and new hospitalizations and ICU admissions reached levels during the second wave of more than double that of the first wave, new deaths in the second wave only marginally surpassed values in the first, and the rate of all three decreased relative to increasing infections. We identify three phases of the probabilities of severe illness reflecting this behavior: the highest values observed during an initial phase from the beginning of the epidemic until May 1, a decreasing trend between May 1 and August 1, and stabilized reduced values following August 1. Specifically, we estimate that between May 1 and August 1, the population-average probability of hospitalization given infection *α*_*t*_ decreased from 0.138 (0.125,0.15) to 0.048 (0.04,0.055); the probability of ICU admission given hospitalization *κ*_*t*_ decreased from 0.597 (0.593,0.603) to 0.543 (0.537,0.549); and the probability of death given ICU admission *δ*_*t*_ decreased from 0.561 (0.557,0.567) to 0.503 (0.49,0.52). The population-average CFR(t) and IFR(t) followed the same trend; we estimate the CFR(t) was 0.0243 (0.021,0.028) on April 15 (corresponding to the first phase values for *α*_*t*_, *κ*_*t*_, and *δ*_*t*_), reached a peak on May 15 of 0.0342 (0.030,0.039), and had dropped to an almost-stable value of 0.0237 (0.02,0.0292) by August 1 (representing the second phase of values for *α*_*t*_, *κ*_*t*_, and *δ*_*t*_), although a slight downward trend still continued until a value of 0.0211 (0.0176,0.0251) on October 15. The IFR(t) was 0.0055 (0.0028,0.0082) on April 15, peaked at 0.0077 (0.0038,0.0115) on May 15, decreased to 0.0055 (0.0024,0.0090) by August 1 and 0.0051 (0.0023,0.0078) by October 15.

### 3.2. Risk-stratified probabilities of severe illness and death for LAC

Table 2 displays the marginal relative risks (RR) extracted from the literature (left column) and conditional RR estimated by the risk model for each risk factor considered by our model on the rates of hospitalization given infection, (*H*|*I*), ICU admission given hospitalization, (*Q*|*H*), and death given ICU admission, (*D*|*Q*). We find that the independent effect of age is stronger than from having any comorbidities. The independent effect of comorbidities and obesity attenuate with increasing severity of disease, while that of age and smoking increase.

Table 1 shows the model-estimated population prevalence, the frequency in the infected population, and the median *P*_*t*_(*H*|*I*), *P*_*t*_(*Q*|*H*), and *P*_*t*_(*D*|*Q*) for each profile with an estimated population prevalence greater than 1% (see appendix p 18 for all profiles). Probabilities are provided for the dates May 15, representing the peak value for the overall CFR(t) and IFR(t) (Figures 3e, 3f), August 1, representing the value of the probabilities after the overall *P*_*t*_(*H*|*I*), *P*_*t*_(*Q*|*H*), and *P*_*t*_(*D*|*Q*) had completed their descent but while the CFR(t) and IFR(t) were still decreasing, and October 15, the most recent date.

**Table 1:**
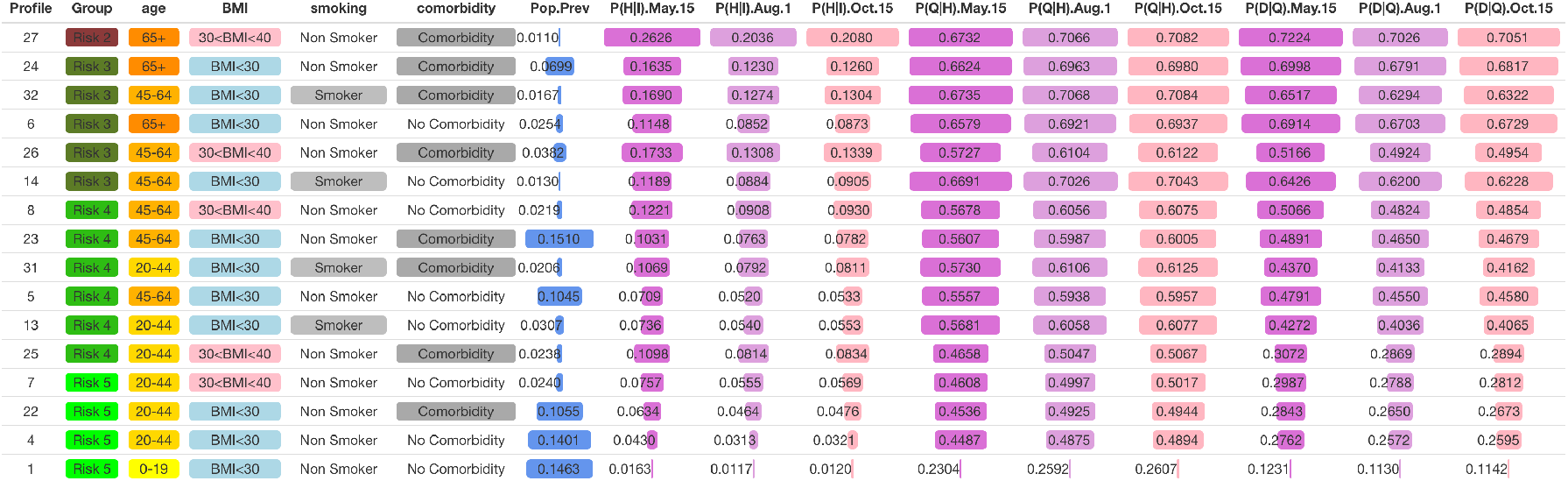
Profile-stratified *P_t_*(*H*|*I*), *P_t_*(*Q*|*H*), and *P_t_*(*D*|*Q*): Risk profiles (characterized by unique combination of age group, BMI range, smoking status, and any comorbidity), risk group (1-5), model-estimated population prevalence, the frequency of each profile in the infected population, and the median probability of hospitalization given infection, ICU admission given hospitalization, and death given admission to the ICU on May 15, August 1, and October 15 2020. Only profiles with a population prevalence *>* 1% are shown.

**Table 2:**
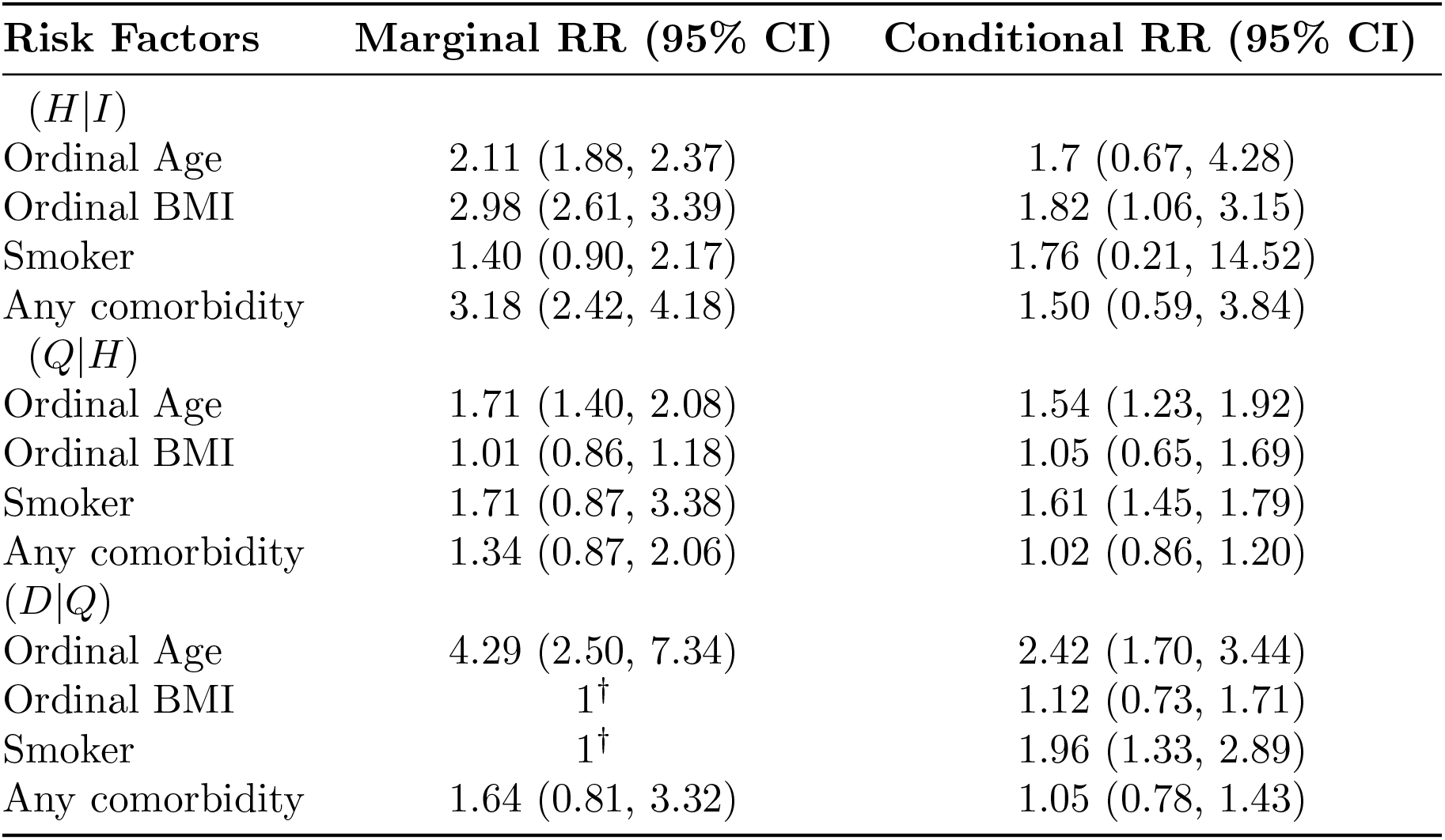
The marginal relative risk collected from published studies on COVID-19 and conditional relative risk estimated by the risk model for each risk factor on rates of hospitalization given infection, (*H*|*I*), ICU admission given hospitalization, (*Q*|*H*) and death given ICU admission, (*D*|*Q*) (95% credible interval). †We set the marginal RR for ordinal BMI and smoker to 1 because we did not find the association between obesity class, smoking status, and the likelihood of death given ICU admission *D*|*Q* due to COVID-19 in the published literature.

| Risk Factors | Marginal RR (95% CI) | Conditional RR (95% CI) |
| --- | --- | --- |
| $(H\vert I)$ | | |
| Ordinal Age | 2.11 (1.88, 2.37) | 1.7 (0.67, 4.28) |
| Ordinal BMI | 2.98 (2.61, 3.39) | 1.82 (1.06, 3.15) |
| Smoker | 1.40 (0.90, 2.17) | 1.76 (0.21, 14.52) |
| Any comorbidity | 3.18 (2.42, 4.18) | 1.50 (0.59, 3.84) |
| $(Q\vert H)$ | | |
| Ordinal Age | 1.71 (1.40, 2.08) | 1.54 (1.23, 1.92) |
| Ordinal BMI | 1.01 (0.86, 1.18) | 1.05 (0.65, 1.69) |
| Smoker | 1.71 (0.87, 3.38) | 1.61 (1.45, 1.79) |
| Any comorbidity | 1.34 (0.87, 2.06) | 1.02 (0.86, 1.20) |
| $(D\vert Q)$ | | |
| Ordinal Age | 4.29 (2.50, 7.34) | 2.42 (1.70, 3.44) |
| Ordinal BMI | 1<sup>†</sup> | 1.12 (0.73, 1.71) |
| Smoker | 1<sup>†</sup> | 1.96 (1.33, 2.89) |
| Any comorbidity | 1.64 (0.81, 3.32) | 1.05 (0.78, 1.43) |

The probability of hospitalization given infection, ICU admission given hospitalization, and death given ICU admission vary extensively across the risk profiles. Notably, the risks within specific marginal factor groups also vary extensively. For example, within the age group 65+, the probability of hospitalization given infection is approximately five times greater for individuals with at least one comorbidity, a smoking history, and severe obesity then for individuals that have no comorbidities, do not smoke, and have a healthy BMI.

### 3.3. Risk-stratified CFR and IFR for LAC

Table 3 shows the frequency in the infected population and the median of the CFR(*t*) and IFR(*t*) on May 15, August 1, and October 15 2020 for each risk profile with a model-estimated population prevalence *>* 1% (see appendix p 20 for all 39 profiles). The probability of death given infection varies tremendously across populations. While the overall median CFR(*t*) ranged from a high of 0.0342 (0.030,0.039) on May 15 to a low of 0.0211 (0.0176,0.0251) on October 15, the profile-stratified median CFR(*t*) ranged from 0.0004 to 0.23 across the profiles. The overall median IFR(*t*) ranged from a high of 0.0077 (0.0038,0.0115) on May 15 to a low of 0.0051 (0.0023,0.0078) on October 15, but ranged from 0.0001 to 0.052 across the profiles. To facilitate interpretation of the probabilities and variability across risk profiles, we group the profiles into five Risk Groups based on similar within-group CFR(*t*) on May 15, with Risk 1 being composed of individuals with CFR(*t*) *>* 0.16; 0.08 *<* CFR(*t*) *<* 0.16 in Risk 2; 0.04 *<* CFR(*t*) *<* 0.08 in Risk 3; 0.01 *<* CFR(*t*) *<* 0.04 in Risk 4; and CFR(*t*) *<* 0.01 in Risk 5 (risk groups are indicated for each profile in Table 3). Notably, profiles including individuals 65+ are found in Groups 1, 2, and 3, and profiles for individuals aged 45-64 are found in Groups 1-4 (appendix p 20).

**Table 3:**
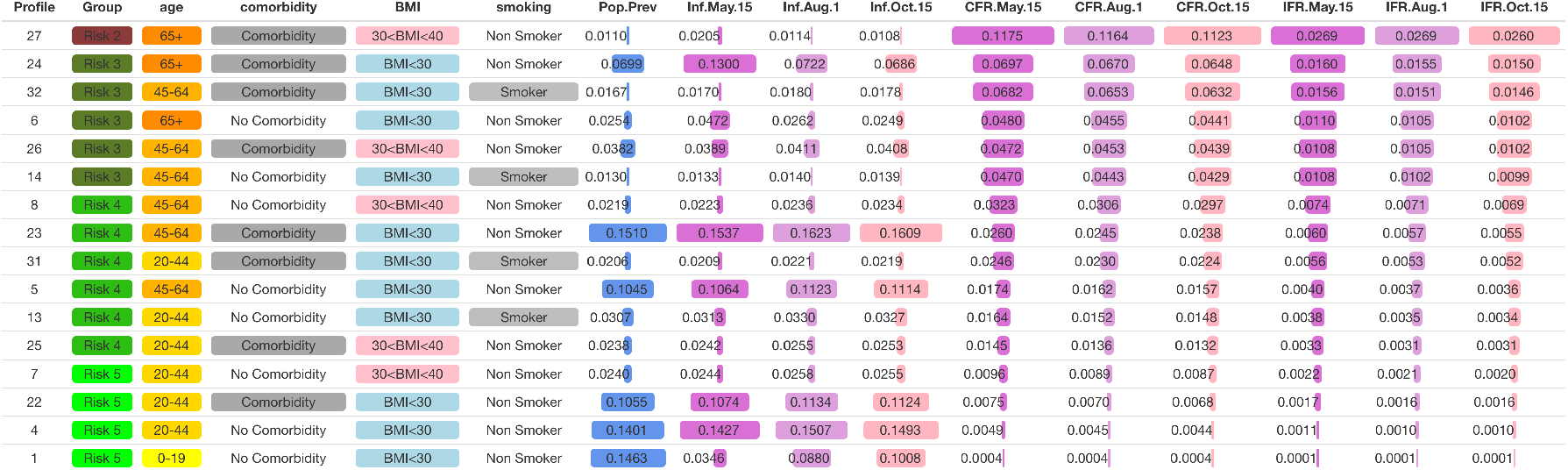
Profile-stratified CFR(*t*) and IFR(*t*): Risk profiles (characterized by unique combination of age group, BMI range, smoking status, and any comorbidity), risk group (1-5), model-estimated population prevalence, the frequency of each profile in the infected population, and the median of the CFR(*t*) and IFR(*t*) on May 15, August 1, and October 15 2020. Only profiles with a population prevalence *>* 1% are shown.

### 3.4. Scenario analysis

Figure 4a shows the three levels of NPI policies implemented (*NPIs=None, Moderate, Observed*) and Figure 4b shows the three levels of isolating the 65+ population implemented (*Protect=Observed, 50%, 100%*) to create the nine scenarios. Figure 5 shows the results of scenarios that include the three *Protect* levels, *NPI=Observed* and *NPI=Moderate*; the results of all scenarios including *NPI=Moderate* are shown in appendix p 22. Results for each scenario by CFR(t), IFR(t), cumulative deaths, current in hospital, current observed infections, and current total infections are pictured. The “Do Nothing” scenario (Scenario 1) results in a median of approximately 75 000 deaths by October 15, exceeding the observed death count in LAC by approximately 11 times the observed median value of 6 855, as well as excessively surpassing hospital capacity for two months with a peak of 40 000 individuals requiring daily hospital care. Combining a stringent protection policy that isolates 100% of individuals 65+ with the “Do Nothing” to reduce community transmission policy (Scenario 3) averts approximately 50% of deaths estimated in the “Do Nothing” Scenario, however surpasses observed deaths in LAC by a factor of 5.5 and exceeds hospital capacity for one month at peak levels 6.5 times as large as maximum capacity. Implementing a moderate NPI policy with no additional measures to protect at-risk populations (Scenario 4) greatly reduces deaths from the scenarios in which no community transmission reduction takes place although still leads to more than 16 000 deaths by October 15, a factor of 2.5 greater than observed.

**Figure 4:**
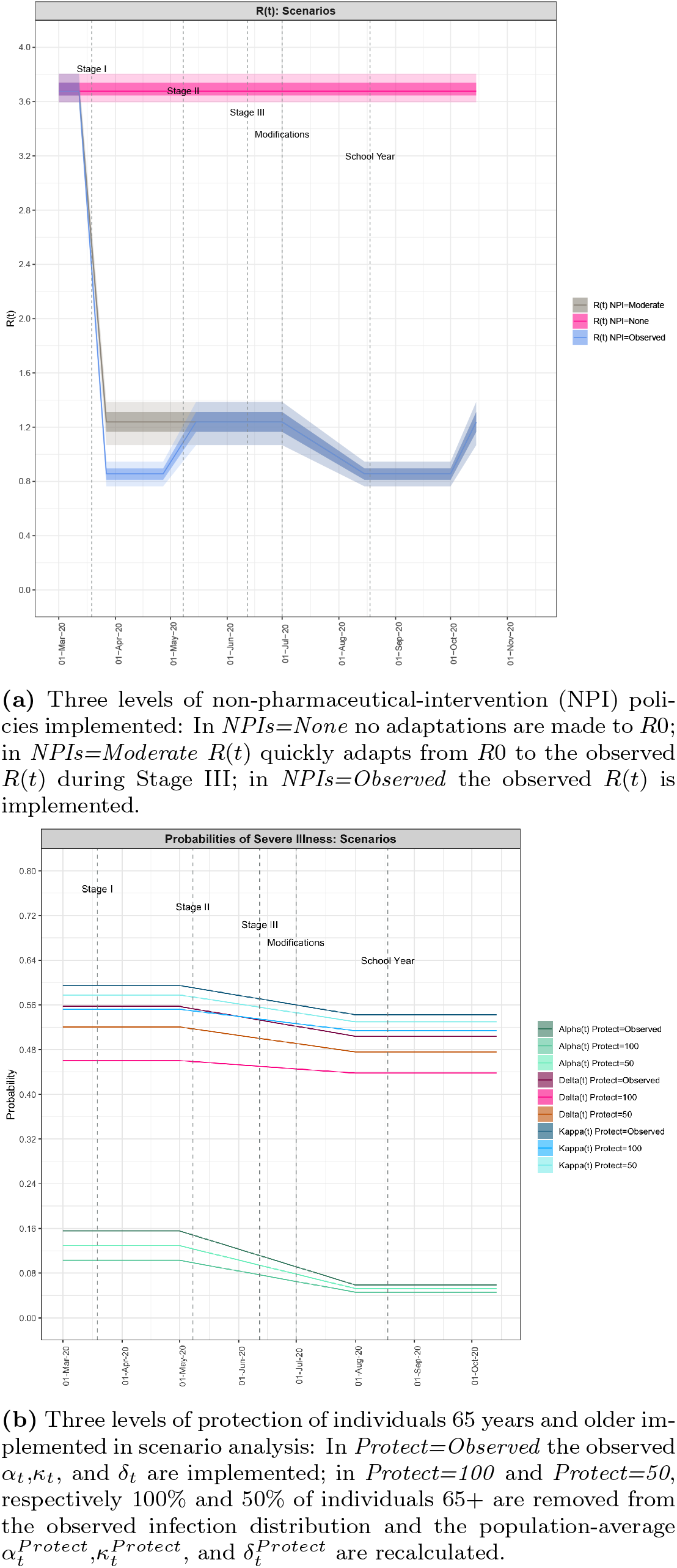
Implemented policies in scenario analysis.

**Figure 5:**
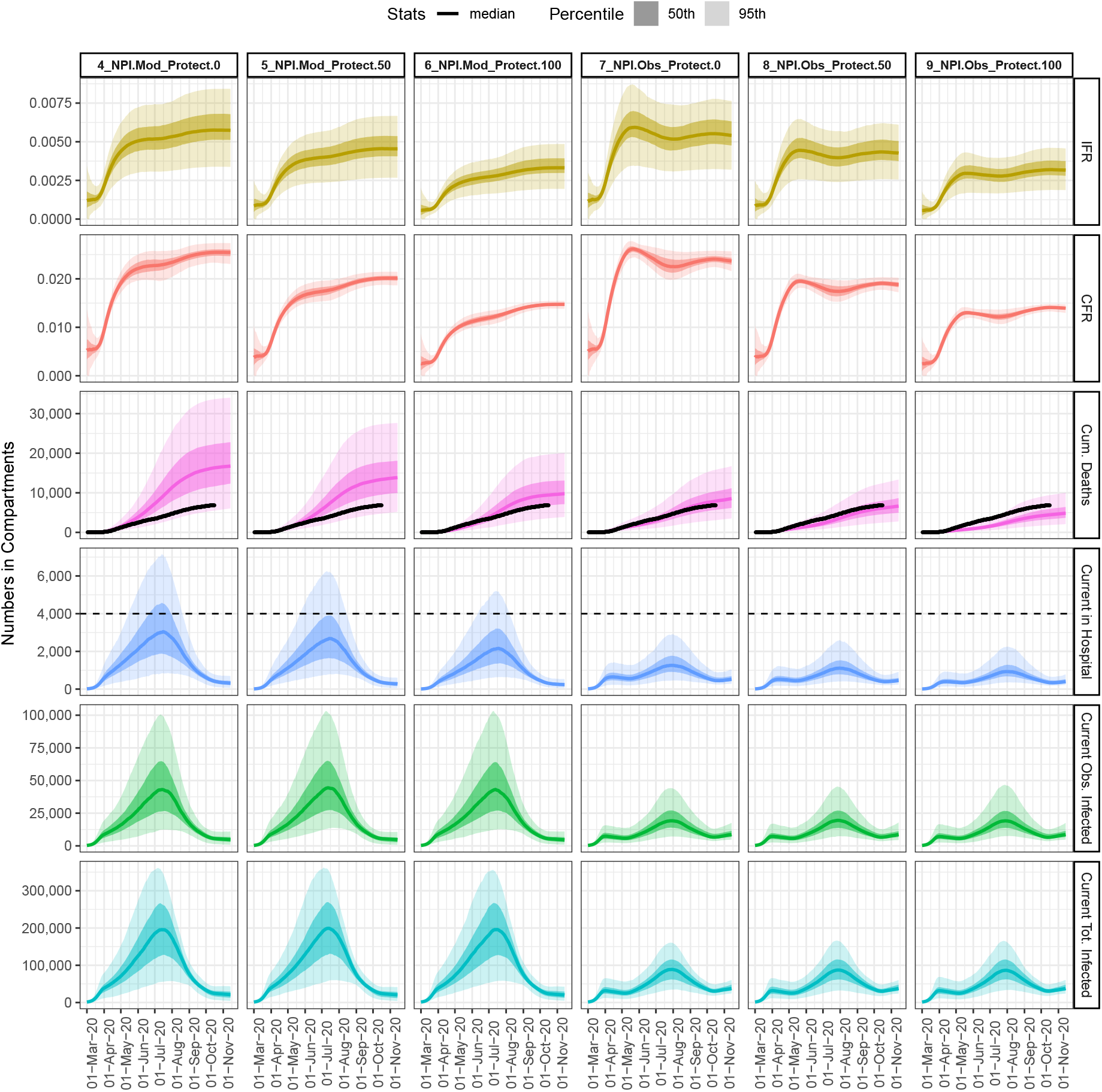
Scenarios implemented and results by CFR(t), IFR(t), cumulative deaths, current in hospital, current observed infections, and current total infections.

When the moderate NPI policy is combined with additional measures to protect 100% of individuals 65+ (Scenario 6), the number of deaths accrued is much closer to the observed death toll in LAC by October 15, while hospital demand would have still approached levels exceeding capacity for two months. If only 50% of individuals 65+ had been protected (Scenario 5), the death count would have doubled the observed count and hospital demand would have come closer to exceeding capacity.

Comparing Scenarios 7-9 in which the observed trend in community transmission is implemented (strict initial lockdown followed by a gradual reopening and stabilization at an *R*(*t*) of approximately 1.26), deaths could have been halved if 100% of individuals 65+ had been protected (Scenario 9) or 1 500 deaths if 50% of individuals 65+ had been protected above the observed self-protection that occurred (Scenario 8). Hospitalization demand would not have decreased appreciably.

The overall population average CFR(t) and IFR(t) are more strongly determined by the implemented *protection* level; by October 15 these approximate and 0.003, respectively, across three scenarios in which 100% of individuals 65+ are protected. Still, much larger absolute numbers of infected individuals, as in the scenario with *NPI=Nothing*, result in much larger absolute numbers of deaths.

## 4. Discussion

This work has developed a framework for using available data on COVID-19 epidemic dynamics and prevalences of COVID-19 risk factors at the population level to estimate time-varying subpopulation-stratified probabilities of severe illness and the case fatality rate (CFR(*t*)) and infection fatality rate (IFR(*t*)) across multiple combinations of risk factors. In the absence of individual-level data, the technical contribution of this work was to integrate a dynamic epidemic model with a risk modeling approach to estimate conditional effects from available marginal data and to subsequently produce time-varying subpopulation-stratified estimates for LAC. To reflect the uncertain knowledge of many parameters and the understanding that in non-linear systems small variations to specific parameters can result in large impacts in outputs [30], we account for uncertainty in all results through the use of a stochastic epidemic model and a Bayesian approach to parameter estimation. On its own, the risk model estimates the conditional effects of each risk factor and therefore the overall effect of risk factors in combination. These adjusted effects have not been typically reported in observational studies on COVID-19, yet help to understand which individuals are at highest risk of advancing to each stage of disease. Integrated together with the epidemic model, the modeling framework allows the analysis of dynamic outcomes and parameters relating to each stage of disease by population and subpopulation.

Analyses demonstrate that the risk of severe illness and death has decreased over time and moreover varies tremendously across subpopulations, suggesting that it is inappropriate to summarize epidemiological parameters for the entire population and epidemic time period. This includes variation not only across age groups, but also within age categories combined with other risk factors analyzed in this study (comorbidities, obesity status, smoking). Across the risk profiles we found median IFRs ranging from 0.0095%-0.04% in the youngest age group (0-19), IFRs ranging from 0.1%-1.8% for those aged 20-44, 0.36%-4.3% for those aged 45-64, and 1.02%-5.42% for those aged 65+ (noting that the 95%CI varied beyond these ranges of median values). The highest IFR for each age strata come from profiles also including comorbidities, obesity Class 2 or 3, and except for the youngest age group, current smoking status.

Our age-stratified IFR are comparable to those found in a recent meta-analysis using results from seroprevalence studies in North America and Europe [8], a model-based analysis for estimating IFR for China [9], and a model-based analysis for estimating IFR in New York City during the time period March 1 through June 6th [10], with the exception that we do not account for further substrata above 65 years of age. This can be seen as a limitation of our analysis of IFR for profiles including age strata above age 65+, since the risk of severe outcomes is known to strongly increase with age. A second limitation of our model-based analysis for older risk strata is that we assume unobserved infections are equally distributed across all risk profiles, whereas there are likely to be far fewer unobserved or asymptomatic infections for those at higher risk of severe outcomes. For risk profiles including individuals age 65+, our IFRs are therefore underestimated and our CFR estimates are likely to be a better approximation of the true IFRs; we find median CFRs in the range 4.4%-23.45% across profiles including individuals 65+, comparable to the range of 2%-22% for IFRs in age groups 65+ found in the meta-analysis [8]. At the same time, the NYC study found IFRs for individuals age 65-74 of 4.87% (3.37%–6.89%), comparable to our median IFR estimate for the 65+ age group, and 14.2% (10.2%–18.1%) for those aged 75 years and older. More generally, in interpreting our results for policy implications, emphasis should be placed on the relative differences in IFR across risk profiles and the understanding that the IFR for a specific age strata represents an average across a wide variation given the presence or absence of other risk factors.

Our overall population-average IFR(*t*) estimates for Los Angeles County, which ranged from a peak of 0.77% (0.38%,1.15%) on May 15 down to 0.55% (0.24%,0.90%) by October 15, are similar to the overall IFR estimated by a model-based analysis for China of 0.66% (95%CI: 0.39%,1.33%), but lower than the overall IFR estimated for NYC as of June 6 of 1.39% (1.04%–1.77%), or 1.10% if only confirmed COVID-19-related deaths were included [10]. This could be explained by the larger outbreak experienced by NYC in the spring, which led to healthcare capacity overload in some parts of the city and larger probabilities of fatality. Our IFR estimates could also be lower since we account only for underascertainment of infections and not of deaths [31, 32]. Even at the lowest overall IFR estimated for LAC as of October 15, a key finding is that COVID-19 is substantially more deadly than seasonal Influenza with a population-average IFR of approximately 0.05% [8, 33].

The time-varying population-average risk probabilities over time provide insights into the changing dynamics of the epidemic in LAC. The decrease in the probability of hospitalization given infection (*α*_*t*_) by almost 200% between the value before May 1 and after August 1 suggests substantive changes in risk of severe illness in the infected population. This follows from the decrease in the prevalence of the age group 65+ in (observed) infections between April 15 and July 15 from approximately 23% to 12% [11], but may also reflect other changes in the demographic composition of infected individuals including other at-risk subpopulations for which LAC data is not available (e.g., individuals with comorbidities), or increases in the prevalence of individuals without access to healthcare [34, 35]. The decrease in the probability of ICU admission given hospitalization (*κ*_*t*_) and death given ICU admission (*δ*_*t*_) between May 1 and August 1 both by approximately 10% suggests a potential effect of better treatment in the healthcare setting [36, 37], as well as seasonal effects [38, 39], but may also result from changes in the infected population making it through to critical illness.

The fact that the risk of severe illness and the CFR(*t*) and IFR(*t*) vary tremendously across populations raises the question of whether protecting certain subpopulations would have achieved similar results in infections, hospitalizations, and deaths as policies applied population-wide to reduce transmission, including the initial lockdown period from March 19 to May 8. We investigate this question through simulation, comparing observed trends with alternate policy scenarios modifying the population-wide transmission rate or protecting specific at-risk subpopulations. We implemented scenarios in which no reductions are made to the transmission rate. However, it is likely that individual behavior would have modified to reduce the transmission rate, as observed in locations where formal shelter-at-home policies were not implemented, such as in Sweden [40]. We therefore implemented a more moderate NPI policy that reduced community transmission at the beginning of the epidemic to levels exhibited during Stage III of the epidemic in LAC, which in practice reflected a panel of public health outreach and restriction policies including mask wearing and closing indoor dining and entertainment, but were not as stringent as the more complete community-wide lockdown of Stage I. We focused on isolating individuals aged 65+ because this was a suggested policy response for the state of California [41], and is the suggested policy plan for initial vaccination distribution [42].

The main finding from the scenario analysis is that the strict initial lockdown period in LAC was effective because it both reduced overall transmission and protected individuals at greater risk, resulting in preventing both healthcare overload and deaths (especially among individuals 65+). Simulation results suggest that an initial implementation of a moderate NPI policy on its own would not have been sufficient to avoid overwhelming healthcare capacity, and additionally would have exceeded the observed death count by close to 2.5 times. Combining the moderate NPI policy with a stringent protection of all individuals 65+ would have resulted in a death count at a similar level as observed, however hospital capacity limits would have approached levels exceeding capacity, likely resulting in more severe illnesses and deaths than accounted for by this model. A more moderate protection scenario in which 50% of individuals 65+ are isolated would have resulted in slightly surpassing the observed death toll and similarly threatening to exceed healthcare capacity. Thus, while similar numbers of deaths as observed in LAC could have been achieved while avoiding initial lockdown through the implementation of a more moderate NPI policy combined with greater protection of individuals 65+, this would have come at the expense of overwhelming the healthcare system. We can therefore conclude that the implemented lockdown was well designed, as in hindsight it appears to have been the minimally required course of action to protect the entire LAC population, and particularly those at high risk of severe disease, while also preventing overcapacity within the healthcare system.

The scenario analysis also has implications for future policy. In anticipation of a continued rise in cases in LAC this winter, policy makers need to consider the trade offs of various policy options on the numbers of the overall population that may become infected, severely ill, and deaths when considering policies targeted at subpopulations at greatest risk of transmitting infection and at greatest risk for developing severe outcomes. Furthermore, this scenario analysis mimics potential vaccination policy related to a population-wide rollout (similar to lockdowns and NPI policies applied to the population) vs. rollout first to specific at-risk populations (similar to scenarios protecting individuals 65+). The results of this scenario analysis suggests policies that first vaccinate only those at greatest risk of severe illness may not be as effective as a more general policy aimed at decreasing the size of the epidemic through vaccinating the overall population; this latter policy, by acting on those at greatest risk of transmitting infection, results in decreasing the total numbers of the population infected as well as subsequent healthcare utilization and deaths [43].

This study is prone to typical limitations occurring when modeling epidemiological dynamics in the context of rapidly evolving infectious disease outbreaks. Data informing the conditional effect estimates within the risk model are aggregated across early, retrospective studies from China (age, comorbidities, smoking) [3] and NYC (BMI) [2] on the fractions of hospitalization, ICU admission, and death by individual risk factors. While we attempt to reframe these results for the demographic composition of the LAC population through regional data on the prevalence of risk factors and the correlation structure between risk factors (appendix p 14), there may be differences in the underlying study population or treatment setting between China, NYC, and LAC that would lead to heterogeneity in effect estimates. However, we believe that the estimates from the Chinese studies do represent population-based estimates as these samples avoid some of the biases present from other potentially available studies, but with highly selected samples.

While this work has focused on demonstrating the substantial heterogeneity in risk probabilities and IFR across subpopulations, we developed a single-population epidemic model. We accounted for differences in the infected population through the observed age distribution in LAC. However, heterogeneity in exposure to COVID-19 infection has been shown to vary extensively across a number of factors including not only age but also race/ethnicity, neighborhood of residence, employment, economic status, and access to PPE, among others [7, 34, 35, 1, 4, 5]. At the time of beginning this study we did not have the data to formally model subpopulation-specific probabilities of exposure or the data on hospitalization and death counts for different groups necessary to fit the parameters of a multi-population model. The approach we developed is a way to use commonly available population-level epidemic timeseries data to model multiple groups in a single population, and combine these population-level estimates with prevalence rates of risk factors to produce stratified estimates for different subpopulations, specific to a given region. Future work will need to develop multi-population models that estimate subpopulation-stratified probabilities and infection rates accounting for key risk factors of both exposure to infection and severe illness given infection. In the meantime, this framework for using limited available data to produce subpopulation-stratified estimates of the severity of illness and CFR/IFR by subpopulations can be generalized to other regional and policy contexts, provided generally-accessible data on epidemic time series and prevalences of marginal risk factors in the overall population.

## Data Availability

All data and code required to reproduce the analysis in this manuscript can be found in the GitHub repository at https://github.com/AbigailHorn/COV2-LA. Specifically, see the `data` folder for all datasets used in our analysis.
Additionally, interactive versions of some of the figures and tables provided in our analysis are provided on the repository website found at https://abigailhorn.github.io/COV2-LA.

## Contributors

ALH and DVC designed the study. ALH wrote the paper with support from DVC. ALH developed the epidemic model and implemented the integrated model. DVC and LJ developed and implemented the conditional risk model. ALH implemented the integrated epidemic and risk model. NS helped to design the policy analysis. ALH led the analysis and visualization of results with assistance from EH. FW prepared the LAC epidemiological data with support from WN and PS. NS, PS, KDH, MP, and WC contributed to writing the paper. DVC and WC coordinated management of the team, including data collection and processing. All authors read and approved the final paper.

NS reported receiving grants from Jedel Foundation, Abbott Diagnostics, the Agency for Healthcare Research and Quality, the National Institutes of Health, the National Institute for Health Care Management Foundation, Health Care Services Corporation, and the Patient-Centered Outcomes Research Institute outside the submitted work; he reported serving as an expert witness for Goldman, Ismail, Tomaselli, Brennan, and Baum; serving as a scientific expert for the American Medical Association, the China Development Research Foundation, and the Pharmaceutical Research and Manufacturers of America; serving as strategic advisor for Payssurance. All authors declare no competing interests.

## Data sharing

For data and code required to reproduce the analysis see https://github.com/AbigailHorn/COV2-LA. The COVID-19 infection, hospitalization, ventilation support, death, and age-stratified infection data analyzed in this paper was provided by the Outbreak Data Coordination Team of the Los Angeles County Department of Public Health, who consented to its sharing.

## Acknowledgements

ALH was supported from an NIH Ruth L Kirschstein National Research Service Award (NRSA) Institutional Training Grant T32 5T32CA009492-35 DVC was supported by NIH P01CA196569 and NCI P30CA014089. EH was supported by NIH P01CA196569. DVC and ALH are also funded by COVID-19 Keck Research Fund from the University of Southern California. We would like to acknowledge data processing and visualization support from Claire Jacquillat.

