## Appendix for "Estimation of COVID-19 risk-stratified epidemiological parameters and policy implications for Los Angeles County through an integrated risk and stochastic epidemiological model"

#### SUPPLEMENTARY APPENDIX

**Abigail L. Horn[1], Lai Jiang[1], Faith Washburn[2], Emil Hvitfeldt[1], Kayla de la Haye[1], William Nicholas[2], Paul Simon[2], Maryann Pentz[1], Wendy Cozen[1], Neeraj Sood[3], & David V. Conti[1]**

[1] **Department of Preventive Medicine**
**Keck School of Medicine**
**University of Southern California**

[2] **Los Angeles County Department of Public Health**
**313 N Figueroa St, Los Angeles, CA 90012**

[3] **Sol Price School of Public Policy**
**University of Southern California**

Basic Reproduction Number, $R0$ • Transmission reduction factor, $\mu(t)$ • Fraction of unreported cases $r(t)$ • Illness severity probabilities • Starting time, $t_0$

2.3 Variable observations used for parameter estimation . . . . . 12

Count variables used in model estimation

2.4 Running the model . . . . . 12

Uncertainty quantification • Model implementation

### II Risk Model 12

**3 Modeled risk factors 13**

**4 Conditional risk effects 13**

Logit transformed population-average probabilities of each stage of disease $\hat{\alpha}_t$, $\hat{\kappa}_t$, $\hat{\delta}_t$ • Vector of conditional log risk estimates for model $m$, $\underline{\hat{\psi}}_m^{Cond}$ • Frequencies of each incoming population $\mathbf{f}_{t,q,in}$ • Mean-centered design matrix $\mathbf{X}_m$, and risk profile design matrix $\mathbf{R}$

6.3 Calculating risk-stratified probabilities . . . . . 17

**7 Resulting profile-stratified probabilities of severe illness and death for LAC 17**

### III Supporting analyses 18

**8 Calculating risk profile stratified CFR and IFR 18**

### Part I
### SEIR+RISK Stochastic Epidemic Model

#### 1 Mathematical Model

We develop a stochastic model of COVID-19 transmission in a single, fully-mixed population. The transmission process we develop reflects transitions known to be important to COVID-19 transmission, including Susceptible, Exposed (latent but not yet infectious), Infectious, and Recovered compartments. We also include healthcare utilization and outcome variables: Hospitalization, Mechanical Ventilation Supported, and Death compartments. This enables us to provide projections for these compartments, which are important for planning. It also enables better specification of our model and more accurate parameter estimation because these data are more reliable than Illness counts. We additionally include a variable and parameter representing the unascertained infectious class. This represents cases that do not appear in the official register of cases, and may include both asymptomatic cases and those with mild symptoms who have not been tested. We estimate this variable by inducing a parameter on the fraction of observed over all cases, with a prior specified on data coming from seroprevalence studies.

##### 1.1 SEIR+Risk variables and parameters

We develop a model of COVID-19 transmission in a single, homogeneously-mixed population divided into 9 compartments representing the number of individuals in a given disease state, or the current "census" in a state. Compartments relating to the transmission of infection are the widely-used susceptible ($S$), exposed ($E$), infectious and observed ($I$), and recovered ($R$). To this we add a compartment representing infectious and unobserved or unascertained ($A$) cases. We model healthcare utilization and outcome at a more granular level by including compartments representing individuals that are hospitalized ($H$), in ICU care ($Q$), undergoing mechanical ventilation support ($V$), and death ($D$). Each individual can only be in one state at each point in time with the exception of $V$, which is a subset of $Q$.

Our model applies the following logic and assumptions. With probability $p_{S\to E}$ (described in the next section), susceptible individuals will become exposed *and* develop infection (emphasizing that exposure to the virus is not a sufficient condition for developing an infection) and move to the exposed but latent state $E$, meaning they will become, but are not yet, infectious. We assume the transfer of susceptible individuals into the exposed state happens at a *per capita* rate $\beta(t)$, defined as the average number of individuals that an infected individual will infect per day. $\beta(t)$, the *transmission rate*, controls the rate of disease spread and reduces following modifications including non-pharmaceutical interventions (NPIs) (Section 1.4).

Including the exposed compartment models the delay between individuals being exposed to infection and becoming infectious. From the exposed and latent state, individuals will transition to one of two active infection states: a time-varying fraction $r(t)$ of these cases will transfer to the observed infectious $I$, and the remaining $1-r(t)$ will transfer to the unobserved infectious state $A$. $I$ represents cases of infection that will eventually test positive for the virus and be confirmed in the official register of infection case data. $A$ represents cases that are symptomatic but do not appear in the confirmed case data (whether because they are asymptomatic, are symptomatic and do not get tested, or get tested and have a false negative result). Therefore, the parameter $r$ represents the fraction of all infectious cases that are observed and confirmed. We assume that individuals transfer from the exposed to infectious and observed ($I$) or unobserved ($A$) compartments at a rate equal to the inverse of the mean latency period, $d_{EI}$.

Infectious cases may either move directly into the recovered state ($R$), or into the healthcare state space if hospitalization ($H$) or further care ($Q$, $V$) are required. Of all observed and infectious cases ($I$), we assume that individuals will require hospitalization with probability $\alpha_t$, equal to $P_t(H|I)$. Infectious individuals transition into the hospitalized state at a rate equal to the inverse of the time between infectiousness and hospitalization, $d_{IH}$, or move directly to recovery at a rate equal to the inverse of the mean time of infection given hospitalization is not required, $d_{IR}$. Hospitalized individuals will require ICU care with a probability $\kappa_t$, equal to $P_t(Q|H)$, and transfer into $Q$ at a rate equal to the inverse of the mean time in hospital given ICU care will be required, $d_{HQ}$. With probability $1-\kappa_t$ they will recover and move into $R$ at a rate of the inverse of the mean time in hospital given ICU care will not be required before recovery, $d_{HR}$. Individuals in ICU may require ventilation support ($V$); we do not model the dynamics into and out of $V$ or represent it as an independent compartment, but rather assume that a fraction $p_V$ of all individuals in ICU will require mechanical ventilation support. We do not include $V$ as an independent state variable because at the time of developing the model we did not have data on the transition times between ICU, ventilation support, recovery, and death, however it was valuable to include $V$ in the model because epidemic timeseries data was available on numbers of individuals receiving ventilation support and not number of individuals in-ICU ($Q$). Individuals in the ICU will recover with probability $1-\delta_t$, moving to R at rate equal to the inverse of the mean time in ICU before recovery and given recovery, $d_{QR}$, or will die with probability $\delta_t$, equal to $P_t(D|Q)$ moving to $D$ with rate equal to the inverse of the mean time in ICU care given a fatal case, $d_{QD}$.

We assume that any individual that enters the healthcare system due to COVID-19 will have a diagnosis, and therefore that all unobserved infected cases ($A$) recover. These cases transition to $R$ at the same rate as observed infectious ($I$) individuals, $1/d_{IR}$. We assume recovered individuals cannot be reinfected due to immunity, and cannot infect others. While the dynamics of transitions within the healthcare setting would change if hospital, ICU, and mechanical ventilator capacity are reached, we do not model this condition as this has not been reached yet in Los Angeles County (LAC). The only route to death is through hospitalization and ICU care. This means we do not model unobserved deaths.

To summarize our assumptions: New infections are created by individuals in the $I$ and $A$ compartments. Individuals in the $E$ compartment are not yet infectious. Individuals in $H$, $Q$, and $V$ are quarantined in the healthcare setting and do not contribute to new infections. Individuals in $R$ and $D$ are removed. Each parameter $d_{IR}, dIH$, etc. represents the mean duration in the previous compartment before transitioning. We differentiate between transition branching probabilities (i.e. $r(t)$, $\alpha_t$, $\kappa_t$, $\delta_t$), which represent the relative probabilities of moving to different compartments, and transition rates ($1/d_{EI}, \dots$), which represent the *per capita* rate at which an individual leaves a compartment. These probabilities and rates are brought together to determine the average transition dynamics between compartments. We model $\beta(t)$, $r(t)$, $\alpha_t$, $\kappa_t$, and $\delta_t$ as varying in time.

State variables are summarized in Table 1. Transition branching probabilities, summarized in Table 3, are estimated by our model while transition rates, summarized in Table 2, are based on values from existing literature. A diagram of the model is shown in Figure 1 in the main text.

##### 1.2 Deterministic differential equation model

The "skeleton" [1] of the compartmental model is described by the following set of coupled ordinary differential equations (ODE) describing the transitions of individuals between the 9 compartments across time:

$$dS/dt = -\beta(t)S(I+A) \tag{1}$$
$$dE/dt = \beta(t)S(I+A) - \tfrac{1}{d_{EI}}E \tag{2}$$
$$dA/dt = \tfrac{1-r(t)}{d_{EI}}E - \tfrac{1}{d_{IR}}A \tag{3}$$
$$dI/dt = \tfrac{r(t)}{d_{EI}}E - \left(\tfrac{\alpha_t}{d_{IH}} + \tfrac{1-\alpha_t}{d_{IR}}\right)I \tag{4}$$
$$dH/dt = \tfrac{\alpha_t}{d_{IH}}I - \left(\tfrac{\kappa_t}{d_{HQ}} + \tfrac{1-\kappa_t}{d_{HR}}\right)H \tag{5}$$
$$dQ/dt = \tfrac{\kappa_t}{d_{HQ}}H - \left(\tfrac{\delta_t}{d_{QD}} + \tfrac{1-\delta_t}{d_{QR}}\right)Q \tag{6}$$
$$dV/dt = p_V Q \tag{7}$$
$$dD/dt = \tfrac{\delta_t}{d_{QD}}Q \tag{8}$$
$$dR/dt = \tfrac{1-\alpha_t}{d_{IR}}I + \tfrac{1-\kappa_t}{d_{HR}}H + \tfrac{1-\delta_t}{d_{QR}}Q + \tfrac{1}{d_{IR}}A \tag{9}$$

With the total population size:

$$P = S+E+A+I+H+Q+D+R \tag{10}$$

where $V$ is not included because it is a sub-set of $Q$.

| Variable | Description |
|---|---|
| $N$ | Total population size |
| $S$ | Susceptible population |
| $E$ | Exposed not yet infectious |
| $A$ | Infected, unobserved |
| $I$ | Infected, observed |
| $H$ | In Hospital |
| $Q$ | In ICU |
| $V$ | On mechanical ventilator |
| $D$ | Dead |
| $R$ | Recovered/removed |

**Table 1.** State variables.

| Parameter | Description | Value | Source |
|---|---|---|---|
| $d_{EI}$ | days between exposure and infectivity (incubation period) | 5 days | [2] |
| $d_{IH}$ | days between symptom onset and hospitalization (if required) | 10 days | [3] |
| $d_{IR}$ | days between symptom onset and recovery (if not hospitalized) | 7 days | [3] |
| $d_{HQ}$ | days between hospitalization and ICU (if required) | 1 day | [4] |
| $d_{HR}$ | days between hospitalization and recovery (if ICU not required) | 12 days | [5] |
| $d_{QD}$ | days between ICU and fatality | 8 days | [5] |
| $d_{QR}$ | days between ICU and recovery | 7 days | [5] |

**Table 2.** Transition rate parameters, values, and sources. These parameters are modeled as fixed values.

| Parameter | Description |
|---|---|
| $R0$ | Basic reproductive number |
| $\beta(t)$ | Time-varying infectivity rate |
| $\mu(t)$ | Time-varying reduction in initial $R0$ |
| $r(t)$ | Time-varying proportion of infections that are detected and reported out of all infections |
| $\alpha_t$ | Time-varying probability infected ($I$) patient requires hospitalization ($H$) |
| $\kappa_t$ | Time-varying probability hospitalized ($H$) patient requires ICU ($Q$) |
| $\delta_t$ | Time-varying probability patient in ICU ($Q$) dies ($D$) |
| $p_V$ | Time-varying probability patient in ICU ($Q$) requires ventilation support ($V$) |

**Table 3.** Transition probability parameters. These parameters are estimated.

##### 1.3 Stochastic dynamical state model

We develop a stochastic discrete-time model of the dynamical state system, which provides an effective framework for parameter estimation and for generating confidence bounds reflecting stochasticity of the disease process in model projections.

We model the number of individuals leaving any of the classes by all available routes over a particular time interval by a set of coupled multinomial counting processes, $N_{X_i \to X_j}(t)$ for all transition pairs $(i, j)$, with random transition rates. Transitions of individuals from one to the next stage of the disease are seen as stochastic movements between the corresponding population compartments at transition rates with random fluctuations. At each period an individual either stays or moves on to the next compartment. It has been proven that in the limit as the time interval $\Delta t \to 0$, the following discrete-time approximation process fulfills the properties defining a continuous time Markov process [6]. We do not model this continuous-time Markov process explicitly and operate only with its discrete approximation.

In general in this process, the random variable for the number of individuals leaving class $X_i$ and going to $X_j$ and $X_k$ over time interval $[t, t+\Delta t)$, $(\Delta N_{X_i \to X_j}(t), \Delta N_{X_i \to X_k}(t))$, has the multinomial distribution

$$(\Delta N_{X_i \to X_j}(t), \Delta N_{X_i \to X_k}(t)) \sim \text{Multinomial}(X_i(t); p_{X_i \to X_j}(t), p_{X_i \to X_k}(t)),$$

for states $X_j$ and $X_k$ reachable from state $X_i$. The multinomial distributions result from summation over individual independent and identical Bernoulli trials for all members of each compartment [7, 8]. If there is only one possible transition direction out of $X_i$, then the distribution above is given by a binomial distribution.

The amount of time spent in a compartment is described by a Poisson process. The time length that an individual spends in a compartment is thus exponentially distributed with some compartment-specific rate $\lambda_i(t) = \lambda_{ij}(t) + \lambda_{ik}(t)$, where $\lambda_{ij}(t)$ and $\lambda_{ik}(t)$ are the rates to go from $X_i$ to $X_j$ and $X_k$, respectively. The probability of extending the stay by a further period of length $\Delta t$ is $\exp(\lambda_i(t)\Delta t)$ and the probability of leaving is $1 - \exp(\lambda_i(t)\Delta t)$ [7]. Given there is a transition out of $X_i$, the conditional probability it is to $X_j$ is $\frac{\lambda_{ij}}{\lambda_{ij}(t)+\lambda_{ik}(t)}$. Therefore the probability of leaving $X_i$ and going to $X_j$ in $[t, t+\Delta t)$, $p_{X_i \to X_j}$, is found as

$$p_{X_i \to X_j} = (1 - \exp(-(\lambda_{ij}(t) + \lambda_{ij}(t))\Delta t)) \frac{\lambda_{ij}}{\lambda_{ij}(t) + \lambda_{ik}(t)}.$$

In the following we proceed to define the increments over $[t, t+\Delta t)$ of all counting process, $\Delta N_{X_i \to X_j}(t)$, defining the transitions between all states in the process.

###### 1.3.1 Counting processes for transitions between states

**New latent infections ($S$ to $E$ transitions)** New infections occur from direct or indirect interactions between susceptible ($S$) and infected ($I$ or $A$) individuals, and arise first in the exposed ($E$) compartment. The rate at which susceptible individuals become infected is described by the *force of infection*, $\Lambda$, which combines the rate of infections per day per infected individual ($\beta(t)$) across all infected individuals by multiplying with the overall fraction of infected individuals at time $t$, $\frac{I(t)+A(t)}{P}$, where $P$ is equal to the total population size; $\Lambda(t) = \beta(t)\frac{I(t)+A(t)}{P}$.

We assume that the corresponding probability at time $t$ that a susceptible individual becomes infected and leaves the $E$ compartment in one time unit $\Delta t$ is equal to $p_{S\to}(t) = 1 - e^{-\beta(t)\frac{I(t)+A(t)}{P}\Delta t}$.

Then, the overall number of individuals becoming infected and moving from the $S$ to the $E$ compartment in one time step, $\Delta N_{S\to E}$, is distributed as a binomial distribution with one draw for each of $S$ individuals, each with probability $p_{S\to E}(t)$, i.e.

$$\Delta N_{S\to E}(t) \sim \text{Binomial}(S(t), p_{S\to E}(t)).$$

**Exposed, latent infections ($E$) becoming infectious ($I$ and $A$)** Exposed and latent individuals will become infectious and transition to the observed infected state ($I$) or the unobserved infected ($A$) state.

The total rate of departures from $E$ is equal to the sum of the rate of departures to $I$, $\frac{r(t)}{d_{EI}}$, and the rate of departures to $A$, $\frac{1-r(t)}{d_{EI}}$, which is simply $\frac{1}{d_{EI}}$. In each time step $\Delta t$, each exposed individual has a time-independent probability of becoming infectious based on this rate, $p_{E\to}(t) = 1 - e^{-\frac{1}{d_{EI}}\Delta t}$. The relative probability that a departure out of $E$ is to $I$ is equal to $r(t)$, and to $A$ is $1 - r(t)$. With these probabilities we can find the number of individuals leaving $E$ and going to $I$ and $A$ over time interval $[t, t+\Delta t)$, $(\Delta N_{E\to I}(t), \Delta N_{E\to A}(t))$, as

$$(\Delta N_{E\to I}(t), \Delta N_{E\to A}(t)) \sim \text{Multinomial}(E(t), (r(t))p_{E\to}(t), (1-r(t))p_{E\to}(t)).$$

**Infection ($I$) to Hospitalization ($H$) or Recovery ($R$)** We model transitions for infectious individuals to recovery ($R$) or hospitalization ($H$).

The total rate of departures out of $I$ is equal to the sum of the rate of departures to $H$, $\frac{\alpha_t}{d_{IH}}$, and the rate of departures to $R$, $\frac{1-\alpha_t}{d_{IR}}$. The probability that an infected individual transitions out of $I$ in each time step is then found as $p_{I\to} = 1 - e^{-(\frac{\alpha_t}{d_{IH}} + \frac{1-\alpha_t}{d_{IR}})\Delta t}$. The relative probability that a departure is to $H$ is equal to $\frac{\frac{\alpha_t}{d_{IH}}}{\frac{\alpha_t}{d_{IH}} + \frac{1-\alpha_t}{d_{IR}}}$, and to $R$ is $\frac{\frac{1-\alpha_t}{d_{IR}}}{\frac{\alpha_t}{d_{IH}} + \frac{1-\alpha_t}{d_{IR}}}$. The number of infected individuals that become hospitalized in each time step, $\Delta N_{I\to H}$, and that recover, $\Delta N_{I\to R}$, are then described by the multinomial distribution,

$$(\Delta N_{I\to H}(t), \Delta N_{I\to R}(t)) \sim \text{Multinomial}(I(t), \frac{\frac{\alpha_t}{d_{IH}}}{\frac{\alpha_t}{d_{IH}} + \frac{1-\alpha_t}{d_{IR}}} p_{I\to}, \frac{\frac{1-\alpha_t}{d_{IR}}}{\frac{\alpha_t}{d_{IH}} + \frac{1-\alpha_t}{d_{IR}}} p_{I\to}).$$

**Transitions out of hospital ($H$) and ICU ($Q$)** We model transitions out of hospital $H$ and out of the ICU $Q$ in the same way as transitions out of $I$, each of which go to two compartments.

The probability of transition in one time step out of $H$ is

$$p_{H\to}(t) = 1 - e^{-(\frac{\kappa_t}{d_{HQ}} + \frac{1-\kappa_t}{d_{HR}})\Delta t}.$$

The random variable for the number of hospitalized individuals that go to the ICU, $\Delta N_{H\to Q}(t)$, and that recover, $N_{H\to R}(t)$, is

$$(\Delta N_{H\to Q}(t), \Delta N_{H\to R}(t)) \sim \text{Multinomial}(H(t), \frac{\frac{\kappa_t}{d_{HQ}}}{\frac{\kappa_t}{d_{HQ}} + \frac{1-\kappa}{d_{HR}}} p_{H\to}(t), \frac{\frac{1-\kappa_t}{d_{HR}}}{\frac{\kappa_t}{d_{HQ}} + \frac{1-\kappa_t}{d_{HR}}} p_{H\to}(t)).$$

The total rate of transitions out of the ICU, to death ($D$) or recovery, is

$$p_{Q\to}(t) = 1 - e^{-(\frac{\delta_t}{d_{QD}} + \frac{1-\delta_t}{d_{QR}})\Delta t}$$

The random variable for the number of individuals in ICU that die, $\Delta N_{Q\to D}(t)$, and that recover, $N_{Q\to R}(t)$, is found as

$$(\Delta N_{Q\to D}(t), \Delta N_{Q\to R}(t)) \sim \text{Multinomial}(Q(t), \frac{\frac{\delta_t}{d_{QD}}}{\frac{\delta_t}{d_{QR}} + \frac{1-\delta_t}{d_{QR}}} p_{Q\to}(t), \frac{\frac{1-\delta_t}{d_{QR}}}{\frac{\delta_t}{d_{QD}} + \frac{1-\delta_t}{d_{QR}}} p_{Q\to}(t)).$$

**Transitions out of unobserved infected state ($A$)** We assume all unobserved infected cases will recover, at the rate $\frac{1}{d_{IR}}$. We model the probability of recovery in one time step as $p_{A\to}(t) = 1 - e^{-\frac{1}{d_{IR}}\Delta t}$. The number of recoveries moving from $A$ to $R$ in each time step is then found as

$$\Delta N_{A\to R}(t) \sim \text{Binomial}(A(t), p_{A\to}(t)).$$

##### 1.3.2 System of equations for state variables and advancing the model

The counting process random variables are coupled to the state variables via the following identities:

$$\begin{aligned}
\Delta S(t) &= -\Delta N_{S\to E}(t)\\
\Delta E(t) &= \Delta N_{S\to E}(t) - \Delta N_{E\to I}(t) - \Delta N_{E\to A}(t)\\
\Delta I(t) &= \Delta N_{E\to I}(t) - \Delta N_{I\to H}(t) - \Delta N_{I\to R}(t)\\
\Delta A(t) &= \Delta N_{E\to A}(t) - \Delta N_{A\to R}(t)\\
\Delta H(t) &= \Delta N_{I\to H}(t) - \Delta N_{H\to Q}(t) - \Delta N_{H\to R}(t)\\
\Delta Q(t) &= \Delta N_{H\to Q}(t) - \Delta N_{Q\to D}(t) - \Delta N_{Q\to R}(t)\\
\Delta V(t) &= p_V \Delta Q(t)\\
\Delta D(t) &= \Delta N_{Q\to D}(t)\\
\Delta R(t) &= \Delta N_{I\to R}(t) + \Delta N_{H\to R}(t) + \Delta N_{Q\to R}(t).
\end{aligned}$$

To simulate from this system we employ an Euler numerical scheme for Markov process compartment models with stochastic rates [6]. We sample the transitions from the counting processes $\Delta N_{X_i\to X_j}(t)$, and update each state variable, representing the number of individuals in each compartment at time $t$, to reflect these counts.

##### 1.3.3 Cumulative and daily counts

The dynamics of the stochastic model are advanced by updating state variables representing the “current census” of individuals in each compartment $X_i$. Observed COVID-19 infection data, however, is available as daily or cumulative counts. We therefore also keep track of auxiliary count variables representing the daily new counts or cumulative counts in specific compartments based on the available data: $I, H, V$, and $R$ (observed data described in Section 2.3).

For cumulative counts we have, based on the counting processes for transitions between compartments already defined:

$$\begin{aligned}
I_{cum}(t+1) &= I_{cum}(t) + \Delta N_{E\to I}(t)\\
H_{cum}(t+1) &= H_{cum}(t) + \Delta N_{I\to H}(t)\\
Q_{cum}(t+1) &= Q_{cum}(t) + \Delta N_{H\to Q}(t)\\
V_{cum}(t+1) &= p_V Q_{cum}(t+1).
\end{aligned}$$

where the $X_{i,cum}$ represents the cumulative version of each state variable $X_i$. $D$ represents a cumulative count and so is not included in the above set of equations. Daily count variables, i.e. $I_{new}, H_{new}, V_{new}$, and $R_{new}$, can be worked out from the cumulative counts as $X_{i,new}(t+1) = X_{i,cum}(t+1) - X_{i,cum}(t)$.

#### 1.4 Time-varying factor reduction in infection rate

We introduce $\mu(t)$, the *transmission reduction factor*. The transmission reduction factor allows us to (i) estimate changes in the transmission rate over time, and (ii) explicitly model changes in the transmission rate with time to simulate different interventions.

The rate at which new infections occur is not constant over the course of an evolving pandemic. The transmission rate $\beta(t)$ can be seen as the product of $p(t)$, the infection probability per contact between an infected and a susceptible individual, and $C(t)$, the individual rate of contacts per time, i.e. $\beta(t) = p(t)C(t)$. Both of these terms may be modified as non pharmaceutical interventions, quarantines, and other policies are introduced to stem spread, in addition to natural modifications in behavior and even evolution in the virulence of the virus strain.

We introduce a time-varying parameter, $\mu(t)$, representing modification to the initial observed value for the infectivity rate parameter, $\beta_0$, from the compounded modifications to $p_0$ and $C_0$ at each time step $t$, i.e.

$$\begin{aligned}
\beta(t) &= \mu(t) p_0 C_0\\
\beta(t) &= \mu(t)\beta_0,
\end{aligned}$$

where $\beta_0$, $p_0$, and $C_0$ represent the initial (averaged) values for the virus in the LAC population before modifications. Therefore $\mu(t)$ and $\beta(t)$ are defined as relative to $\beta_0$.

With $\mu(t)$, the *force of infection* is modified as $\tilde{\Lambda} = \mu(t)\beta_0 \frac{I(t)+A(t)}{P}$, and the corresponding overall number of individuals becoming infected and moving from the $S$ to the $E$ compartment in one time step is modified as

$$\Delta\tilde{N}_{S\to E}(t) \sim \text{Binomial}(S(t), \tilde{p}_{S\to E}(t)),$$

where $\tilde{p}_{S\to E}(t) = 1 - e^{-\mu(t)\beta_0 \frac{I(t)+A(t)}{N(t)}\Delta t}$.

$\mu(t)$ can be specified according to any functional form. In this work we apply a pieceweise linear function (see Section 2.2.2).

##### 1.5 Solving for the basic reproduction number, $R_0$

The basic reproductive number, $R0$, is a dimensionless parameter defined as the average number of secondary infections produced by a single infected individual during that individual's entire period of infection, assuming a completely susceptible population [9]. While not a parameter of the system of state variables, $R0$ is a parameter of interest because it provides a measure of the likelihood that an epidemic will occur within a population.

In a deterministic model, $R0$ represents an epidemic threshold for which values of $R0 < 1$ indicate a lack of disease spread, and values of $R0 > 1$ are consistent with epidemic spread. For a stochastic model, if $R0 > 1$ the probability that an epidemic will develop is less than 1, and if $R0 < 1$, extinction of the infection will occur with a probability less than 1. On average, however, no epidemic will occur if $R0 \leq 1$ [10]. Therefore, estimates of $R0$ provide clearly communicable insight regarding the degree of intensity of interventions required to achieve control [9].

For a simple SIR model, $R0$ is a simple expression equal to the product of the number of infections per time for an infected individual, i.e. the transmission rate $\beta$, and the average duration of the infectious period, e.g. $d_I$, that is,

$$R0 = \frac{\text{infections}}{\text{time}} \times \frac{\text{time}}{\text{infection}} = \beta d_I.$$

Because our model involves multiple infectious classes with different rates of infection duration, the average number of individuals an infected individual infects is a more complex expression. We use the *Next Generation Matrix* (NGM) [11] approach to find the expression for $R0$. The NGM approach involves computing $R0$ by linearizing the ODE model in Section 1.2 around the infection-free steady state and identifying conditions that guarantee increases in infected states [11]. We therefore focus on the *infection subsystem*, i.e. the subset of ODEs that describe the production of new infections and changes in the states of already existing infections only, which is

$$\begin{aligned} dE/dt &= \beta_0 S(I+A) - \tfrac{1}{d_{EI}}E \\ dA/dt &= \tfrac{1-r_0}{d_{EI}}E - \tfrac{1}{d_{IR}}A \\ dI/dt &= \tfrac{r_0}{d_{EI}}E - (\tfrac{\alpha_0}{d_{IH}} + \tfrac{1-\alpha_0}{d_{IR}})I, \end{aligned}$$

where $r_0$ and $\alpha_0$ represent the initial estimated values of these parameters.

Setting $X = (\frac{dE}{dt}, \frac{dA}{dt}, \frac{dI}{dt})^\intercal$, we write the linearized infection subsystem in the form

$$\frac{dX}{dt} = (T + \Sigma)X,$$

where $T$ corresponds to all epidemiological events leading to new infections, and $\Sigma$ corresponds to all epidemiological events relating to transitions in or out of these infected states. We write the new infection matrix $T$ as

$$T = \begin{pmatrix} 0 & \beta_0 & \beta_0 \\ 0 & 0 & 0 \\ 0 & 0 & 0 \end{pmatrix}$$

and the new transmission matrix $\Sigma$ as

$$\Sigma = \begin{pmatrix} -\frac{1}{d_{EI}} & 0 & 0 \\ \frac{r_0}{d_{EI}} & -(\frac{\alpha_0}{d_{IH}} + \frac{1-\alpha_0}{d_{IR}}) & 0 \\ \frac{1-r_0}{d_{EI}} & 0 & -\frac{1}{d_{IR}} \end{pmatrix}.$$

Diekmann et al. [11] show that $R0$ can be found as the dominant eigenvalue of the next generation matrix $K_L = -T\Sigma^{-1}$, or equivalently, the dominant eigenvalue of $K = E^{\intercal}\Sigma^{-1}E$ , where $E$ is an auxiliary matrix whose columns consist of unit vectors relating to the non-zero rows of $T$. $E$, which in this case is equal to $E^{\intercal} = \begin{bmatrix} 1 & 0 & 0 \end{bmatrix}$ , is included to reduce dimensionality.

We find $\Sigma^{-1}$ as

$$\Sigma^{-1} = \begin{pmatrix} d_{EI} & 0 & 0 \\ \frac{-r_0}{\frac{\alpha_0}{d_{IH}} + \frac{1-\alpha_0}{d_{IR}}} & \frac{1}{\frac{\alpha_0}{d_{IH}} + \frac{1-\alpha_0}{d_{IR}}} & 0 \\ -(1-r_0)d_{IR} & 0 & d_{IR} \end{pmatrix}.$$

We have $E^{\intercal}T = \begin{bmatrix} 0 & \beta_0 & \beta_0 \end{bmatrix}$, and $\Sigma^{-1}E = \begin{bmatrix} d_{EI} & \frac{-r_0}{\frac{\alpha_0}{d_{IH}} + \frac{1-\alpha_0}{d_{IR}}} & -(1-r_0)d_{IR} \end{bmatrix}^{\intercal}$. We can then find the next generation matrix as,

$$K = -E^{\intercal}T\Sigma^{-1}E = \beta_0 \left[ \frac{-r_0}{\frac{\alpha_0}{d_{IH}} + \frac{1-\alpha_0}{d_{IR}}} + (1-r_0)d_{IR} \right].$$

$R0$ is the dominant eigenvalue of $K$, which is simply equal to $K$ in this case. Therefore,

$$R0 = \beta_0 \left[ \frac{r_0}{\frac{\alpha_0}{d_{IH}} + \frac{1-\alpha_0}{d_{IR}}} + (1-r_0)d_{IR} \right].$$

We note that the NGM approach for deriving $R0$ is based on the deterministic ODE equations, while we use this parameter with the stochastic model. We justify deriving it from the deterministic model because for large population sizes $N$, the mean of the stochastic equations will converge on the deterministic model equations [10, 12].

We see that $R0$ is directly proportional to $\beta_0$. If $R0$ is the value of the reproductive number at the beginning of the pandemic period before modifications, we define the time-varying reproductive number as $R(t)$. Modifications to the infectivity rate as $\beta(t) = \mu(t)\beta_0$ can be interpreted as equivalent to modifications to $R(t)$, i.e. $R(t) = \mu(t)R0$.

#### 2 Parameter Estimation

Transmission rate parameters we model as fixed values taken from the literature (Table 2). The remaining six model parameters, $\theta = \{\beta(t), r(t), \alpha_t, \kappa_t, \delta_t, p_V\}$ (Table 3), must be estimated with data. To this set of six parameters we add an additional parameter, $t_0$, representing the starting time of the epidemic in LAC; that is, the very first case of COVID-19, which may have been unobserved. This parameter links the epidemic model to the observed count data based on calendar dates. In this section we describe how we estimate these parameters using approximate Bayesian computation (ABC) techniques, COVID-19 count data from the Los Angeles County Department of Public Health, and auxiliary data informing prior parameter specification.

##### 2.1 Parameter estimation through approximate Bayesian computation (ABC)

We use Approximate Bayesian Computation Markov chain Monte Carlo (ABC-MCMC) techniques to estimate the parameters of our mathematical model from available data [13, 14]. ABC provides a number of benefits that make it particularly well suited to this application, allowing estimation of model parameters when: (1) a likelihood function is intractable or unknown; (2) multiple data features are available for model estimation, e.g. infection, hospitalization, and death counts; (3) data is missing, partially observed, or uncertain, such as unreliable early infection surveillance data; (4) prior information and/or assumptions are available about the distribution or range of values each parameter may take [15]. MCMC applying the Metropolis-Hastings algorithm helps to efficiently guide the approach into promising regions of the parameter space [13, 14].

#### 2.2 Specifying prior parameter distributions

##### 2.2.1 Basic Reproduction Number, $R0$

While $R0$ is not a parameter of the epidemic model, it is directly related to the parameter $\beta_0$ through Equation 1.5. Because prior estimates of $R0$ are more readily available in the existing literature than estimates for $\beta_0$, we estimate $R0$, from which we then obtain an estimate for $\beta_0$. $R0$ depends on other model parameters: the fixed parameters $d_{IH}$ and $d_{IR}$, and the estimated parameters $\alpha_t$ and $r(t)$. Therefore $R0$, $\alpha_t$, and $r(t)$ are estimated jointly in the ABC framework.

We base the prior distribution for $R0$ on values for $R0$ estimated from other published studies on COVID-19. A review of $R0$ values estimated for COVID-19 across studies involving different locations, time periods, and modeling approaches reported values in the range of [1.40-6.49], with a mean of 3.28 [16]. Values for $R0$ in SEIR models are also higher than those in $SIR$ models, as has been demonstrated by previous researchers [17]; for example, Bertozzi et al. [18] demonstrate an estimated $R0$ for California of 2.4 using a SIR model and 4.9 using an SEIR model. We therefore model $R0$ according to a normal distribution centered around 3.5, with a standard deviation chosen such that 95% of the area of the distribution is within approximately $\pm 0.5$ of the mean. The modeled prior distribution for $R0$ is therefore:

$$R0 \sim N(3.5, 0.25)$$

##### 2.2.2 Transmission reduction factor, $\mu(t)$

Geolocation trace data from smartphones, i.e. mobility data, can be used estimate changes in distances travelled, commuting, time spent outside of home, visits to specific types of venues, and encounter rates in the community [19]. Because of the relation $\beta(t) = \mu(t) p_0 C_0$ (see Section 1.4), reductions in contact rate $C_0$ correspond to proportional reductions in $\beta(t)$, and equivalently, $R(t)$.

We use mobility data to inform the magnitude and the timing of inflection points in the transmission reduction factor $\mu(t)$. We reference mobility data for LAC from a dashboard provided by Unacast, a company that builds and maintains anonymized mobility datasets based on geolocation data aggregated from millions of users across the U.S. [20]. Unacast provides three metrics: the *Change in Average Mobility (Based on Distance Traveled)*, the *Change in Non-Essential Visits*, and the *Difference in Encounter Density*. We reference the metrics *Change in Average Mobility (Based on Distance Traveled)* and *Difference in Encounter Density* (Figure 1) because these both inform the changes and timing of changes in contact rates, in different ways [19].

Across these two metrics we observe the following trends in $\mu(t)$: (i) a sharp initial descent from the original contact rate beginning around March 12 2020 (a few days before county-wide quarantine policy was announced on March 15), and leveling out around March 28; (ii) a steady plateau during the initial period of the lockdown from around March 28 - April 26; (iii) a steady incline in contact rates from April 27 - July 1 (when measures to close societal businesses were re-announced); (iv) a lower plateau beginning August 15 (when schools opened), and (v) an increase beginning October 1. The increase in October is not modeled on the mobility metrics, but is required for the model to achieve accurate fits to data; this likely reflects the fact that $\mu(t)$ represents changes to both the contact rate and the probability of transmission and may include decreasing NPI adherence, more time spend indoors, and other conditions that may increase transmissiblility of the virus.

To model these trends, we develop a distribution with the change points in time given in Table 4 and two estimated values, $\mu_1$ and $\mu_2$, reflecting values of $\beta(t)$ (and $R(t)$) that are toggled between. A piecewise linear function is interpolated between dates.

This modeled function implies that the estimated $R0$, as in the function $R0(t) = \mu(t)R0$, applies to the time interval from $t_0 \leq t$ <March 12.

##### 2.2.3 Fraction of unreported cases $r(t)$

The prior distribution for $r(t)$, the fraction of infections that are observed, is estimated on April 15 ($r_1$), and August 15, 2020 ($r_2$). We interpolate a linear change between the values on the two dates, and assume that $r(t)$ before April 15 is equal to $r_1$ and after August 15 is equal to $r_2$, i.e. the distribution shown in Table 5. The prior for $r_1$ is allowed to vary over an interval truncated by minimum and maximum values calculated by seroprevalence studies performed

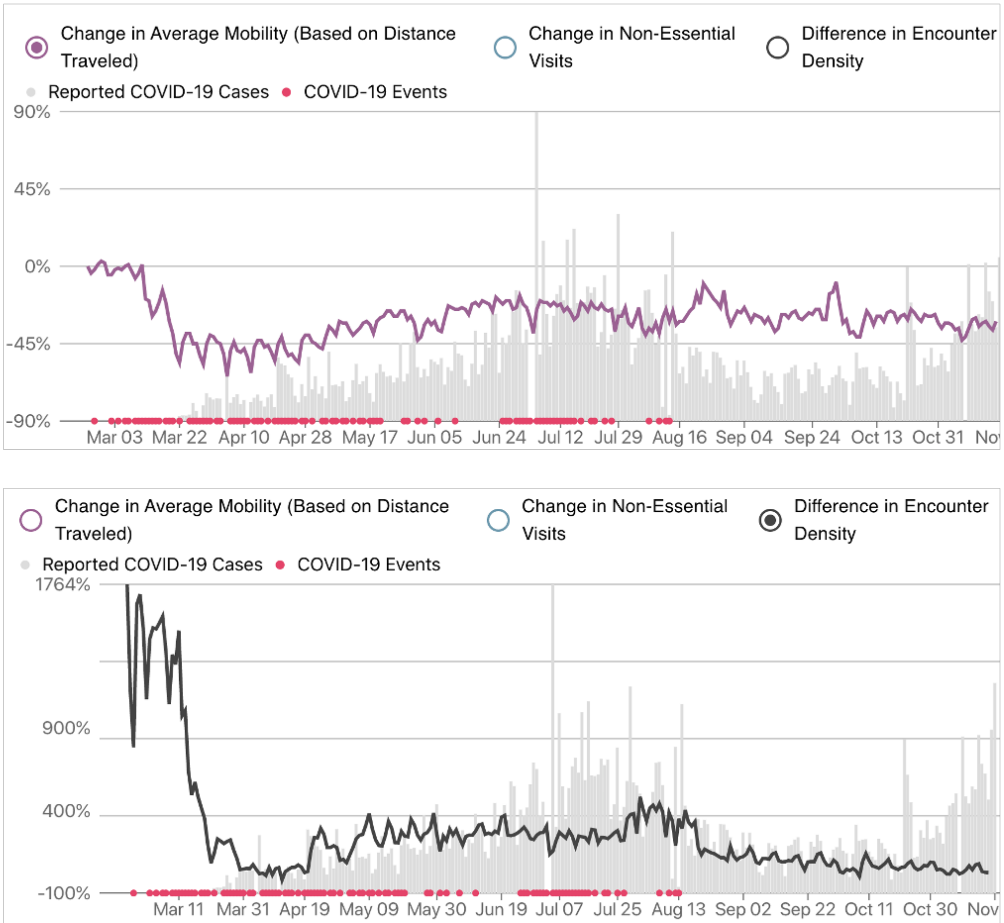

**Figure 1.** The *Change in Average Mobility (Based on Distance Traveled)* and the *Difference in Encounter Density* metric for Los Angeles County from March 1 - Nov 15, 2020 from the the Unacast dashboard [20].

| **Date** | $\mu(\mathbf{t})$ | **Prior Distribution** |
|---|---|---|
| 2020-03-01 | $\mu_0$ | 1 |
| 2020-03-12 | $\mu_0$ | 1 |
| 2020-03-27 | $\mu_1$ | $\sim \mathrm{Unif}(0.2, 0.05)$ |
| 2020-04-27 | $\mu_1$ | $\sim \mathrm{Unif}(0.2, 0.05)$ |
| 2020-05-15 | $\mu_2$ | $\sim \mathrm{Unif}(0.33, 0.05)$ |
| 2020-07-01 | $\mu_2$ | $\sim \mathrm{Unif}(0.33, 0.05)$ |
| 2020-08-15 | $\mu_1$ | $\sim \mathrm{Unif}(0.2, 0.05)$ |
| 2020-10-01 | $\mu_1$ | $\sim \mathrm{Unif}(0.2, 0.05)$ |
| 2020-10-15 | $\mu_2$ | $\sim \mathrm{Unif}(0.33, 0.05)$ |

**Table 4.** Prior distribution for the transmission reduction factor $\mu(t)$ modeled on mobility data from Unacast [20].

| date | Prior $r(t)$ |
|---|---|
| 2020-03-01 | $\sim$ Unif$(0.175, .145)$ |
| 2020-04-15 | $\sim$ Unif$(0.175, .145)$ |
| 2020-08-15 | $\sim$ Unif$(0.4, 0.35)$ |
| 2020-10-15 | $\sim$ Unif$(0.4, 0.35)$ |

**Table 5.** Prior distribution for the fraction of observed cases over all infections, $r(t)$.

| date | Prior $\alpha\_t$ | Prior $\kappa\_t$ | Prior $\delta\_t$ |
|---|---|---|---|
| 2020-03-01 | $\sim$ N$(0.14, 0.01)$ | $\sim$ N$(0.6, 0.01)$ | $\sim$ N$(0.55, 0.01)$ |
| 2020-04-30 | $\sim$ N$(0.14, 0.01)$ | $\sim$ N$(0.6, 0.01)$ | $\sim$ N$(0.55, 0.01)$ |
| 2020-05-01 | $\sim$ N$(0.14, 0.01)$ | $\sim$ N$(0.6, 0.01)$ | $\sim$ N$(0.55, 0.01)$ |
| 2020-08-01 | $\sim$ N$(0.05, 0.01)$ | $\sim$ N$(0.55, 0.01)$ | $\sim$ N$(0.52, 0.01)$ |
| 2020-10-15 | $\sim$ N$(0.05, 0.01)$ | $\sim$ N$(0.55, 0.01)$ | $\sim$ N$(0.52, 0.01)$ |

**Table 6.** Prior distributions for the time-varying probabilities of each disease stage over time: probability of hospitalization given infection, $\alpha_t$; probability of ICU admission given hospitalization, $\kappa_t$; probability of death given ICU admission, $\delta_t$.

by the CDC in March - early May, which were found to be as few as 34.7 times as many infections as observed cases, and as many as 3.2 times as many cases observed as estimated, in the study regions [21]. We allow the second date $r(t)$ to vary over a much wider interval since we do not have prior information corresponding to this time period.

###### 2.2.4 Illness severity probabilities

We describe how we estimate the time-varying probabilities of each disease stage over time $\alpha_t$, $\kappa_t$, and $\delta_t$, which we estimate at two values with a linear function interpolated between (see Figure 3b in the main text).

Changes in these probabilities are determined by the changing risk profile of the infected population. The only available infection data for LAC by risk factor was for age [22]. We therefore used the distribution of infections by age to inform the timescale and shape of the linear function modeling $\alpha_t$, $\kappa_t$, and $\delta_t$ over time. From the beginning of the epidemic in LAC in early March through mid-April, the fraction of infections made up by individuals 65 years and above (*65+*) increased to approximately 23% and the fraction of infections made up by individuals aged 0-19 remained stable at around 1-2%. From April 15 until July 15, the trend in the infection distribution by age reversed, with individuals 65+ decreasing to 12% and individuals 0-19 increasing to 9%. From July 15 through mid-October this relative age distribution remained relatively stable [22].

We assume this distribution results in three phases in the values representing $\alpha_t$, $\kappa_t$, and $\delta_t$: Initial values before the decrease began, a linear decrease in values between beginning the beginning and end of the decrease, and stabilized values following the decrease. There is a delay in the effect of the changing profile of infections by age on the population-average probabilities of severe illness and death, as the infected population advances across the stages of disease. We therefore model a corresponding change in $\alpha_t$, $\kappa_t$, and $\delta_t$ across the date range May 1 - August 1, 2020. The full linear distribution can be modeled by estimating $\alpha_t$, $\kappa_t$, and $\delta_t$ on May 1 and August 1, i.e. the distribution provided in Table 6. Prior distributions for each probability on each date were modeled as normal distributions with means informed by the ratios of observed numbers of illnesses, hospitalizations, and deaths in LAC on each date.

###### 2.2.5 Starting time, $t_0$

The first observed case of COVID-19 in LAC is registered as occurring on January 25 2020, and the epidemic began to grow exponentially in early March 2020 [23]. It is likely that the epidemic in LAC had multiple starting points. We therefore model $t_0$ as a uniform distribution with the unobserved first initial case beginning anywhere between January 1 and February 15 2020.

#### 2.3 Variable observations used for parameter estimation

The model was fit to the daily and cumulative count of infected, hospitalized, undergoing ventilation support, and dead individuals in LAC from a data set from the LACDPH, updated daily by the COVID-19 Outbreak Data Coordination Team and shared privately [22], with a few modifications. Similar count data for LAC, without the jurisdictions of Long Beach and Pasadena, which are included in the privately shared data, can be found on the LACDPH COVID-19 dashboard [23].

##### 2.3.1 Count variables used in model estimation

We use the time series of counts in the compartments Infected (observed) $(I)$, Hospitalized $(H)$, Mechanical Ventilation $(V)$, Deaths $(D)$, and Recovered $(R)$ in the parameter estimation procedure. For each state variable, we use both its daily count, $X_{i,new}(t)$, and its cumulative count, $X_{i,cum}(t)$, representations. We truncate the data such that we do not include infection observations from early in the epidemic, when observations are more likely to be incomplete and unreliable. Specifically, we compute the summary statistic, $Summ(D)$, of the data $D$: $Summ(D) = \{I(t > \text{March 15}, H(t > \text{March 20}), V(t > \text{March 20}), D(t > \text{March 25}), R(t_0 < t < \text{April 4})\}$.

#### 2.4 Running the model

##### 2.4.1 Uncertainty quantification

We use ABC to estimate all model parameters simultaneously, producing joint posterior probability estimates over all parameters. In forward simulations of our model, we simulate trajectories with parameter values coming from this joint posterior distribution, rather than a single value of a parameter. Uncertainty is also contributed by the stochastic differential equations, which model stochasticity in numbers of transitions between compartments.

To produce uncertainty estimates, we estimate credible intervals (CIs) for all model variables and estimated parameters by quantifying uncertainty from two sources: variability due to joint estimated parameter values, and variability due to the stochastic variability between model runs with the same parameters. We aggregate simulations from 1000 jointly estimated parameter sets. For each parameter set, we aggregate simulations for 500 stochastic epidemic model realizations. We pool together all simulations and report their median and 2.5th/97.5th percentiles.

##### 2.4.2 Model implementation

The model was implemented in R (version 3.6.3).

**Epidemic model initial values** In our experiments, $t = 0$ is the ABC-estimated value for $t_0$, which is linked to a calendar date, and will vary across the estimated parameter sets. We set $S(0)$ is equal to 10 million, approximately the population of LAC; $E(0) = 1$; and all other compartments initially empty.

**Parameter estimation implementation details** We apply the R package EasyABC [24] and the algorithm *Marjoram*, which implements the algorithm of Marjoram et al. (2003) [13] with improvement steps by Wegmann et al. (2009) [14]. The arguments that need to be specified are the prior parameter distributions, the original data $D$, the epidemic model to simulate data $D^*$ from, and the function that computes summary statistics $Summ(D)$. The additional parameters that need to be specified are the number $n$ of simulations to perform in the calibration step; the number of parameter values to accept $N$, the *tolerance quantile* $\eta$ that helps to determine the tolerance level $\xi$; and the *scale factor* $\phi$ that helps to determine the size of the proposal range.

We set $n = 100,000$ burn-in simulations, $N = 1000$ sets of jointly estimated parameter values to save, and use the default values for tolerance quantile of $\eta = 0.01$ and scale factor of $\phi = 1$. We test the model for convergence by running the parameter estimation procedure multiple times with different seeds and verifying similarity across variable outputs. Prior parameter distributions were adjusted such that convergence is achieved.

### Part II
### Risk Model

The *risk model* produces estimates, stratified across 39 combinations of modeled risk factors (i.e. *risk profiles*), of the probability of each stage of disease given infection within LAC. These profile-stratified estimates are produced by integrating population-average estimates from the epidemic model with estimates of the conditional relative risk of each modeled risk factor, (age, existing comorbidities, obesity, smoking) and of the prevalence of each *risk profile* in the infected population in LAC.

First, we use the epidemic model to estimate the population-average probability that individuals in LAC who acquire infection are admitted to hospital, $\hat{\alpha}_t$, who are in hospital require admittance to the ICU, $\hat{\kappa}_t$, and who are in ICU will die, $\hat{\delta}_t$ (Section 2.2.4). Second, we calculate conditional risk effects estimates for three models ($m$): (1) hospitalization given illness, $(H|I)$, (2) ICU admission given hospitalization, $(Q|H)$, and (3) death given hospitalization, $(D|Q)$, using available data from published studies on the marginal effects of individual risk factors (age, existing comorbidities, obesity, smoking) (Section 4). Third, we estimate the prevalence of each risk profile in the infected population using available data on the prevalence of each marginal risk factor in the LAC population and the frequency of each marginal age group over infections in LAC (Section 5). Finally, we integrate the probability estimates from the epidemic model $\hat{\alpha}_t$, $\hat{\kappa}_t$, and $\hat{\delta}_t$ with the corresponding conditional risk estimates for each model $(H|I)$, $(Q|H)$, and $(D|Q)$, and the prevalence of each risk profile in the infected population to produce estimates for the disease stage probabilities across all risk profiles, $\underline{\widehat{P_t(H|I)}}$, $\underline{\widehat{P_t(Q|H)}}$, and $\underline{\widehat{P_t(H|I)}}$ (Section 6). In the paper we report the probability estimates at three time points: May 15, August 1, and October 15, 2020.

#### 3 Modeled risk factors

The risk factors, $p$, included in our conditional estimate analysis are:

- **Age**
- **Body mass index (BMI)**
- **Smoker**
- **Other comorbidities**: diabetes, hypertension, chronic obstructive pulmonary disease (COPD), hepatitis B, coronary heart disease, stroke, cancer and chronic kidney disease.

We modeled age and BMI as an ordinal variable and assume an additive effect of both age and BMI on the three outcomes. Age was categorized within four groups: $0-19$, $20-44$, $45-64$, and $65+$, and BMI was categorized in three groups according to obesity classes: Class 1 (no obesity) $BMI < 30\frac{kg}{m^2}$; Class 2 (obesity), $30 \leq BMI \leq 40\frac{kg}{m^2}$; Class 3 (severe obesity), $BMI > 40\frac{kg}{m^2}$. Any comorbidity and smoking status were modeled as binary variables. Note that risk factors are age, BMI, smoking and comorbidities, but age has 4 categories and BMI has 3 categories so $p = \{1,..,9\}$.

#### 4 Conditional risk effects

We estimate the conditional relative risk (RR) effects corresponding to each risk factor for each of the three risk models using marginal effects estimates available from reported studies and a method called the joint analysis of marginal summary statistics (JAM) [25]. JAM uses two pieces of information: (i) the marginal effect estimates between risk factors and outcomes for each model, and (ii) a reference correlation structure between the risk factors. For information informing (i) we obtain the log marginal RR between individual risk factors $p$ and COVID-19 infection severity for each model $m$, $\psi_{p,m}^{Marg}$, from peer-reviewed clinical studies on patients with laboratory-confirmed COVID-19 [26, 27]. For (ii), we obtain the reference correlation structure, $\Sigma$, using data from The National Health and Nutrition Examination Survey (NHANES) from 2017-2018 [28].

##### 4.1 Marginal effect estimates between risk factors from published literature

We extracted the marginal RRs for each risk factor from clinical studies on patients with laboratory-confirmed COVID-19. Ordinal age, smoker, and any comorbidity we extracted from [26], a study reporting outcomes for 1099 patients from 552 hospitals in 30 provinces, autonomous regions, and municipalities in mainland China. The marginal RR of ordinal BMI were extracted from [27], with 4103 COVID-19 patients with laboratory-confirmed COVID-19 treated at a single academic health system in New York City. The left column of the main text Table 2 in the main text displays the marginal RRs extracted from the literature (95% confidence interval), $\psi_{p,m}^{Marg}$, for each risk factor and each of the three models hospitalization given illness, $(H|I)$, ICU admission given hospitalization, $(Q|H)$ and death given ICU admission, $(D|Q)$.

##### 4.2 Correlation structure between risk factors

We obtain the correlation structure, $\Sigma$, between the risk factors $p$ using data from The National Health and Nutrition Examination Survey (NHANES) [28]. NHANES is a survey research program conducted by the National Center for Health Statistics to assess the health and nutritional status of adults and children in the United States, and to track changes over time. We use the NHANES cohort of 2017-2018. To make the correlation matrix representative of the LAC population, we calculated it separately for each race/ethnicity and weight the correlation matrix by the distribution of the race/ethnicity in LA County. The resulting correlation structure $\Sigma$ between the four risk factor categories (age, comorbidities, BMI, smoking) is shown in Figure 2.

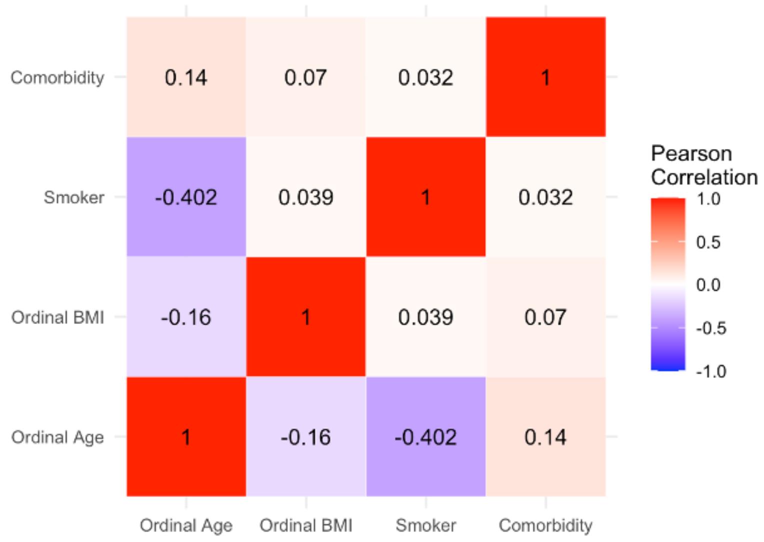

**Figure 2.** Weighted correlation structure of the four risk factors: ordinal age, ordinal BMI, smoker, and any comorbidity, based on data from NHANES [28].

##### 4.3 JAM conditional estimates and risk profiles

We apply the JAM methodology to calculate the conditional log relative risk (RR) for each risk factor, $\psi_p^{Cond}$, from the log of the marginal RR $\psi_p^{Marg}$ and the correlation structure between risk factors $\Sigma$. The *reference group* was set as patients aged $0-19$, $BMI \leq 30\ \frac{kg}{m^2}$, non-smoker, and no comorbidities. See [25] for details on the statistical methodology. The resulting relative risk effects (i.e., $\exp \hat{\psi}_{p,m}^{Cond}$) calculated by JAM for each risk factor and each of the three models are shown in the right column of Table 2 in the main text.

#### 5 Frequency of each risk profile in the infected population in LAC

We estimate the time-varying frequency of each risk profile in the infected population, $\mathbf{f}_{t,q,I}$. First, we use available LAC data the prevalence of the individual risk factors within the overall LAC population [29] [30] to estimate the frequency of each risk profile within the overall population. Second, we use available data on the prevalence of each age group in illnesses [22] together with our estimate of the prevalence of each risk factor in the overall LAC population to estimate the frequency of each profile within the infected population on each date. Illness timeseries data by age group is used because this is the only individual risk factor with observed infection prevalence data in LAC.

#### 5.1 Frequency of risk profiles in overall LAC population

While data exists on the prevalence of single risk factors in the LAC population, data does not exist on the prevalence of the multi-factorial risk profiles $q$ that hold significance for risk of severe COVID-19 illness. We therefore estimate the frequency of the risk profiles $q$ in the overall LAC population, $l_q$, by simulating a sample population based on the prevalence of each individual risk factor in LAC and the weighted correlation structure between the risk factors obtained from NHANES data, $\Sigma$ (Section 4.2).

##### 5.1.1 Data sources

The prevalence of ordinal age and smoking are taken from the Los Angeles County Health Survey (LACHS), study year 2018 [29]. To construct the obesity variable we find the $BMI \leq 30\frac{kg}{m^2}$ and $BMI > 30\frac{kg}{m^2}$ classes from LACHS. We then divide the $BMI > 30\frac{kg}{m^2}$ into the $30 \leq BMI < 40\frac{kg}{m^2}$ and $BMI > 40\frac{kg}{m^2}$ classes based on the relative ratios between these classes on average across the U.S., in data from the Behavioral Risk Factor Surveillance System study year 2010 [31]. The prevalence of cancer comes from the California Health Information Survey (CHIS) [30].

The prevalence of each of the 9 risk factors are combined in the vector $\mathbf{l}_p$.

##### 5.1.2 Estimation

To calculate the frequency of the risk profiles $q$ in the overall LAC population, $l_q$, we first generate a simulated population $\chi$ by sampling from a multivariate normal, $\chi \sim N(x; \mathbf{l}_p, \Sigma)$, where $x$ is the number of samples, $\mathbf{l}_p$ is the vector of the prevalence of each individual risk factor in LAC, and $\Sigma$ is the correlation structure between the risk factors as described in Section 4.2. Each sample from $\chi$ represents a specific risk profile, sampled in proportion to the probability of the combination of risk factors co-occurring. Prevalence of the *any comorbidity* category is constructed by sampling profiles expanded across all comorbidities, and creating an aggregated binary *any comorbidity* variable that takes value 1 if any of the comorbidities are included in the sampled profile.

We then calculate the vector of the frequencies of each risk profile $q$ in the overall LAC population, $\mathbf{l}_q$, as its relative frequency in the simulated population $\chi$.

#### 5.2 Frequency of risk profiles in infected population

We obtain the frequency of each age group over illnesses, $f_{t,p',I}$ from the daily LACDPH data digest [22]. To estimate the frequency of each risk profile $q$ in the infected population, $f_{t,q,I}$, the frequency of each age group over infections is then stratified across the risk profiles according to the relative frequency of each profile within each age group in the overall LAC population.

We find $\mathbf{f}_{t,q,I}$, the vector of the frequency of each risk profile in the infected population, as follows.

##### 5.2.1 Profile design matrix

First we define a risk profile design matrix $\mathbf{R}$, a $q$ $x$ $p$ matrix, that includes indicator variables for the presence or absence of each risk factor $p$ (columns) within each risk profile $q$ (row), encoding the linear combination of all (plausible) linear combinations of the risk factors:

$$\mathbf{R} = \begin{bmatrix} R_{11} & \dots & R_{q1} \\ \dots & \dots & \dots \\ R_{1p} & \dots & R_{qp} \end{bmatrix}.$$

We also define $\mathbf{R}_{p'}$ to be a profile design matrix encoding age group risk factors only, i.e., a $q$ $x$ 4 matrix representing the first four columns of $\mathbf{R}$.

##### 5.2.2 Calculation

We find the vector of the frequency of the risk profiles in the infected population, $\mathbf{f}_{t,q,I}$, using $\mathbf{l}_q$, $\mathbf{f}_{t,p',I}$, and $\mathbf{R}_{p'}$.

First, let $\mathbf{l}_{p'}$ be a vector representing the frequency of each age group in the (estimated) overall population, found as $\mathbf{l}_{p'} = \mathbf{l}_q^{\intercal}\mathbf{R}_{p'}$.

Then we find $\mathbf{f}_{t,q,I}$ as

$$\mathbf{f}_{t,q,I} = \mathbf{l}_q \cdot \frac{\mathbf{R}_{p'}\mathbf{f}_{t,p',I}}{\mathbf{R}_{p'}\mathbf{l}_{p'}}.$$

### 6 Integrated risk model

#### 6.1 Overview

We estimate the time-varying disease stage probabilities stratified across all risk profiles, $q$, for the three models $m=1: P_t(H|I)$, $m=2: P_t(Q|H)$, and $m=3: P_t(D|Q)$. This is done by combining in a logistic regression framework all (plausible) linear combinations of the $p$ risk factors specified in a mean-centered design matrix, $\mathbf{X}_m$, and their corresponding conditional log RR obtained from JAM, $\hat{\psi}^{Cond}_{p,m}$ (Section 4), with intercepts that are set to the logit of the estimated probabilities from the epidemic model for $\hat{\alpha}_t$, $\hat{\kappa}_t$, $\hat{\delta}_t$ (Section 2.2.4), respectively. The matrix $\mathbf{X}_m$ is a profile design matrix that has been mean-centered on the frequency of each risk profile in the incoming population relevant to each model $m$; namely, the infected ($I$) population for model 1, the hospitalized ($H$) population for model 2, and the in-ICU ($Q$) population for model 3. The specification of the frequency of each risk profile in the infected ($I$) population is described in Section 5. The frequency of each risk profile in the hospitalized population, $\mathbf{f}_{t,q,H}$ and the in-ICU population, $\mathbf{f}_{t,q,Q}$, are calculated recursively from the estimated frequency of each profile in the incoming population to each stage of disease, described in Section 6.2.3. Patients age $20-44$ with the risk factors non-smoker, $BMI < 30\frac{kg}{m^2}$, and no comorbidity are assumed to be the reference profile.

We produce estimates for the disease stage probabilities across all risk profiles, $\underline{\widehat{P_t(H|I)}}$, $\underline{\widehat{P_t(Q|H)}}$, and $\underline{\widehat{P_t(H|I)}}$, corresponding to estimates for $\hat{\alpha}_t$, $\hat{\kappa}_t$, and $\hat{\delta}_t$ and the frequency of each profile over infections $\mathbf{f}_{q,I}$ on the same dates.

#### 6.2 Inputs

##### 6.2.1 Logit transformed population-average probabilities of each stage of disease $\hat{\alpha}_t$, $\hat{\kappa}_t$, $\hat{\delta}_t$

In Section 2.2.4 we estimate the population-average probability that individuals in LAC who acquire infection are admitted to hospital, $\hat{\alpha}_t$, who are in hospital require admittance to the ICU, $\hat{\kappa}_t$, and who are in ICU will die, $\hat{\delta}_t$. We logit transform these probabilities as

$$\text{logit}(\hat{\alpha}_t) = \log(\frac{\hat{\alpha}_t}{1-\hat{\alpha}_t})$$
$$\text{logit}(\hat{\kappa}_t) = \log(\frac{\hat{\kappa}_t}{1-\hat{\kappa}_t})$$
$$\text{logit}(\hat{\delta}_t) = \log(\frac{\hat{\delta}_t}{1-\hat{\delta}_t}).$$

##### 6.2.2 Vector of conditional log risk estimates for model $m$, $\underline{\hat{\psi}}^{Cond}_m$

In Section 4 we described the conditional log risk effects estimates, i.e., $\hat{\psi}^{Cond}_{p,m}$ for risk factor $p$ and model $m$. We bring the conditional log risk effect estimates for model $m$ together in to the vector $\underline{\hat{\psi}}^{Cond}_m$.

##### 6.2.3 Frequencies of each incoming population $\mathbf{f}_{t,q,in}$

In Section 5 we estimated the frequency of each risk profile $q$ in the infected population, $\mathbf{f}_{t,q,I}$ (a vector). The frequency of each risk profile in the hospitalized population, $\mathbf{f}_{t,q,H}$ and the in-ICU population, $\mathbf{f}_{t,q,Q}$, are calculated recursively from the estimated frequency of each profile in the incoming population to each stage of disease, as the normalized product of the frequency in the incoming stage of disease and the probability of advancing to the subsequent stage, normalized over all risk profiles.

Specifically, the frequency of each risk profile in the hospitalized population, $\mathbf{f}_{t,q,H}$, is calculated from the estimated $\widehat{P_t(H|I)}$ and $\mathbf{f}_{t,q,I}$ as:

$$\mathbf{f}_{t,q,H} = \frac{\widehat{\mathbf{P_t(H|I)}} \cdot \mathbf{f_{t,q,I}}}{\widehat{\mathbf{P_t(H|I)}}^{\mathsf{T}} \mathbf{f_{t,q,I}}},$$

and the frequency of each risk profile in the in-ICU population, $\mathbf{f}_{t,q,Q}$, is calculated from the estimated $\widehat{P_t(Q|H)}$ and $\mathbf{f}_{t,q,H}$ as:

$$\mathbf{f}_{t,q,Q} = \frac{\widehat{\mathbf{P_t(Q|H)}} \cdot \mathbf{f_{t,q,H}}}{\widehat{\mathbf{P_t(Q|H)}}^{\mathsf{T}} \mathbf{f_{t,q,H}}}.$$

In the following we will require the marginal frequency of each risk factor $p$ in each incoming population, $\mathbf{f}_{t,p,I}$, which we find by marginalizing the risk factors over the risk profiles $q$ for each population as $\mathbf{f}_{t,p,in} = \mathbf{f}^{\mathsf{T}}_{t,q,in}\mathbf{R}$.

###### 6.2.4 Mean-centered design matrix $\mathbf{X}_m$, and risk profile design matrix $\mathbf{R}$

The mean-centered design matrix for model $m$, $\mathbf{X}_m$, is a profile design matrix that has been mean-centered on the frequency of each risk profile in the incoming population $m_{in}$ relevant to each model $m$, $\mathbf{f}_{p,m_{in}}$; namely, the infected ($I$) population for model 1, the hospitalized ($H$) population for model 2, and the in-ICU ($Q$) population for model 3.

We find the mean-centered design matrices as:

$$\mathbf{X}_1 = \mathbf{R} - \mathbf{f}_{t,p,I}$$
$$\mathbf{X}_2 = \mathbf{R} - \mathbf{f}_{t,p,H}$$
$$\mathbf{X}_3 = \mathbf{R} - \mathbf{f}_{t,p,Q}.$$

##### 6.3 Calculating risk-stratified probabilities

We find the probability of each risk profile for model 1, $\underline{\widehat{P_t(H|I)}}$, using the frequency of each risk factor in the infected population, $\mathbf{f}_{p,I}$, as:

$$\underline{\widehat{P_t(H|I)}} = \text{expit}(\text{logit}(\hat{\alpha}_t) + \mathbf{X_1}\underline{\hat{\psi}}^{Cond}_{m=1})$$

We can then find the probability of each risk profile for model 2, $\underline{\widehat{P_t(Q|H)}}$, using the frequency of each risk factor in the hospitalized population, $\mathbf{f}_{p,H}$, as:

$$\underline{\widehat{P_t(Q|H)}} = \text{expit}(\text{logit}(\hat{\kappa}_t) + \mathbf{X_2}\underline{\hat{\psi}}^{Cond}_{m=2}).$$

Finally, we find the probability of each risk profile for model 3, $\underline{\widehat{P_t(D|Q)}}$, using the frequency of each risk factor in the in-ICU population, $\mathbf{f}_{p,Q}$, as:

$$\underline{\widehat{P_t(D|Q)}} = \text{expit}(\text{logit}(\hat{\delta}_t) + \mathbf{X_3}\underline{\hat{\psi}}^{Cond}_{m=3}).$$

#### 7 Resulting profile-stratified probabilities of severe illness and death for LAC

The resulting probabilities across each risk profile for each of three models $\underline{\widehat{P_t(H|I)}}$, $\underline{\widehat{P_t(Q|H)}}$, and $\underline{\widehat{P_t(D|Q)}}$, as well as the estimated frequency of each profile in the overall LAC population and the infected population, on the dates May 15, August 1, and October 15, 2020, are shown in Table 7.

To facilitate interpretation of the probabilities and variability across risk profiles, we group the risks into 5 groups based on similar within-group CFR($t$) on May 15 (Figure 8), with Risk 1 being composed of individuals

| Profile | Group | age | BMI | smoking | comorbidity | Pop.Prev | P(H\|I).May.15 | P(H\|I).Aug.1 | P(H\|I).Oct.15 | P(Q\|H).May.15 | P(Q\|H).Aug.1 | P(Q\|H).Oct.15 | P(D\|Q).May.15 | P(D\|Q).Aug.1 | P(D\|Q).Oct.15 |
|---|---|---|---|---|---|---|---|---|---|---|---|---|---|---|---|
| 36 | Risk 1 | 65+ | 30<BMI<40 | Smoker | Comorbidity | 0.0002 | 0.3863 | 0.3113 | 0.3171 | 0.7690 | 0.7956 | 0.7968 | 0.8357 | 0.8220 | 0.8237 |
| 30 | Risk 1 | 65+ | BMI>40 | Non Smoker | Comorbidity | 0.0026 | 0.3935 | 0.3178 | 0.3236 | 0.6839 | 0.7166 | 0.7182 | 0.7458 | 0.7270 | 0.7294 |
| 38 | Risk 1 | 45-64 | BMI>40 | Smoker | Comorbidity | 0.0005 | 0.4031 | 0.3265 | 0.3324 | 0.6946 | 0.7267 | 0.7282 | 0.7019 | 0.6813 | 0.6838 |
| 18 | Risk 1 | 65+ | 30<BMI<40 | Smoker | No Comorbidity | 0.0001 | 0.2947 | 0.2308 | 0.2356 | 0.7655 | 0.7923 | 0.7936 | 0.8301 | 0.8160 | 0.8178 |
| 33 | Risk 2 | 65+ | BMI<30 | Smoker | Comorbidity | 0.0019 | 0.2568 | 0.1988 | 0.2031 | 0.7602 | 0.7875 | 0.7888 | 0.8200 | 0.8053 | 0.8071 |
| 12 | Risk 2 | 65+ | BMI>40 | Non Smoker | No Comorbidity | 0.0009 | 0.3010 | 0.2361 | 0.2410 | 0.6796 | 0.7126 | 0.7142 | 0.7382 | 0.7190 | 0.7214 |
| 20 | Risk 2 | 45-64 | BMI>40 | Smoker | No Comorbidity | 0.0003 | 0.3094 | 0.2434 | 0.2484 | 0.6904 | 0.7227 | 0.7243 | 0.6935 | 0.6725 | 0.6751 |
| 27 | Risk 2 | 65+ | 30<BMI<40 | Non Smoker | Comorbidity | 0.0110 | 0.2626 | 0.2036 | 0.2080 | 0.6732 | 0.7066 | 0.7082 | 0.7224 | 0.7026 | 0.7051 |
| 35 | Risk 2 | 45-64 | 30<BMI<40 | Smoker | Comorbidity | 0.0039 | 0.2704 | 0.2102 | 0.2146 | 0.6842 | 0.7168 | 0.7184 | 0.6762 | 0.6547 | 0.6574 |
| 15 | Risk 2 | 65+ | BMI<30 | Smoker | No Comorbidity | 0.0005 | 0.1865 | 0.1414 | 0.1446 | 0.7566 | 0.7841 | 0.7854 | 0.8140 | 0.7989 | 0.8008 |
| 9 | Risk 2 | 65+ | 30<BMI<40 | Non Smoker | No Comorbidity | 0.0037 | 0.1911 | 0.1451 | 0.1484 | 0.6688 | 0.7024 | 0.7040 | 0.7143 | 0.6942 | 0.6967 |
| 17 | Risk 2 | 45-64 | 30<BMI<40 | Smoker | No Comorbidity | 0.0017 | 0.1974 | 0.1501 | 0.1535 | 0.6798 | 0.7128 | 0.7144 | 0.6674 | 0.6456 | 0.6483 |
| 29 | Risk 2 | 45-64 | BMI>40 | Non Smoker | Comorbidity | 0.0090 | 0.2763 | 0.2152 | 0.2197 | 0.5847 | 0.6220 | 0.6238 | 0.5465 | 0.5224 | 0.5254 |
| 37 | Risk 3 | 20-44 | BMI>40 | Smoker | Comorbidity | 0.0013 | 0.2844 | 0.2220 | 0.2267 | 0.5967 | 0.6336 | 0.6354 | 0.4941 | 0.4700 | 0.4729 |
| 24 | Risk 3 | 65+ | BMI<30 | Non Smoker | Comorbidity | 0.0699 | 0.1635 | 0.1230 | 0.1260 | 0.6624 | 0.6963 | 0.6980 | 0.6998 | 0.6791 | 0.6817 |
| 32 | Risk 3 | 45-64 | BMI<30 | Smoker | Comorbidity | 0.0167 | 0.1690 | 0.1274 | 0.1304 | 0.6735 | 0.7068 | 0.7084 | 0.6517 | 0.6294 | 0.6322 |
| 11 | Risk 3 | 45-64 | BMI>40 | Non Smoker | No Comorbidity | 0.0044 | 0.2022 | 0.1540 | 0.1575 | 0.5798 | 0.6172 | 0.6191 | 0.5366 | 0.5124 | 0.5154 |
| 19 | Risk 3 | 20-44 | BMI>40 | Smoker | No Comorbidity | 0.0007 | 0.2087 | 0.1592 | 0.1628 | 0.5919 | 0.6290 | 0.6308 | 0.4841 | 0.4600 | 0.4629 |
| 6 | Risk 3 | 65+ | BMI<30 | Non Smoker | No Comorbidity | 0.0254 | 0.1148 | 0.0852 | 0.0873 | 0.6579 | 0.6921 | 0.6937 | 0.6914 | 0.6703 | 0.6729 |
| 26 | Risk 3 | 45-64 | 30<BMI<40 | Non Smoker | Comorbidity | 0.0382 | 0.1733 | 0.1308 | 0.1339 | 0.5727 | 0.6104 | 0.6122 | 0.5166 | 0.4924 | 0.4954 |
| 14 | Risk 3 | 45-64 | BMI<30 | Smoker | No Comorbidity | 0.0130 | 0.1189 | 0.0884 | 0.0905 | 0.6691 | 0.7026 | 0.7043 | 0.6426 | 0.6200 | 0.6228 |
| 34 | Risk 3 | 20-44 | 30<BMI<40 | Smoker | Comorbidity | 0.0044 | 0.1791 | 0.1354 | 0.1386 | 0.5849 | 0.6222 | 0.6240 | 0.4642 | 0.4402 | 0.4431 |
| 8 | Risk 4 | 45-64 | 30<BMI<40 | Non Smoker | No Comorbidity | 0.0219 | 0.1221 | 0.0908 | 0.0930 | 0.5678 | 0.6056 | 0.6075 | 0.5066 | 0.4824 | 0.4854 |
| 16 | Risk 4 | 20-44 | 30<BMI<40 | Smoker | No Comorbidity | 0.0053 | 0.1265 | 0.0942 | 0.0965 | 0.5800 | 0.6175 | 0.6193 | 0.4543 | 0.4304 | 0.4333 |
| 28 | Risk 4 | 20-44 | BMI>40 | Non Smoker | Comorbidity | 0.0070 | 0.1835 | 0.1390 | 0.1422 | 0.4780 | 0.5170 | 0.5189 | 0.3333 | 0.3121 | 0.3146 |
| 23 | Risk 4 | 45-64 | BMI<30 | Non Smoker | Comorbidity | 0.1510 | 0.1031 | 0.0763 | 0.0782 | 0.5607 | 0.5987 | 0.6005 | 0.4891 | 0.4650 | 0.4679 |
| 31 | Risk 4 | 20-44 | BMI<30 | Smoker | Comorbidity | 0.0206 | 0.1069 | 0.0792 | 0.0811 | 0.5730 | 0.6106 | 0.6125 | 0.4370 | 0.4133 | 0.4162 |
| 10 | Risk 4 | 20-44 | BMI>40 | Non Smoker | No Comorbidity | 0.0073 | 0.1298 | 0.0967 | 0.0991 | 0.4730 | 0.5120 | 0.5139 | 0.3244 | 0.3036 | 0.3061 |
| 5 | Risk 4 | 45-64 | BMI<30 | Non Smoker | No Comorbidity | 0.1045 | 0.0709 | 0.0520 | 0.0533 | 0.5557 | 0.5938 | 0.5957 | 0.4791 | 0.4550 | 0.4580 |
| 13 | Risk 4 | 20-44 | BMI<30 | Smoker | No Comorbidity | 0.0307 | 0.0736 | 0.0540 | 0.0553 | 0.5681 | 0.6058 | 0.6077 | 0.4272 | 0.4036 | 0.4065 |
| 25 | Risk 4 | 20-44 | 30<BMI<40 | Non Smoker | Comorbidity | 0.0238 | 0.1098 | 0.0814 | 0.0834 | 0.4658 | 0.5047 | 0.5067 | 0.3072 | 0.2869 | 0.2894 |
| 7 | Risk 5 | 20-44 | 30<BMI<40 | Non Smoker | No Comorbidity | 0.0240 | 0.0757 | 0.0555 | 0.0569 | 0.4608 | 0.4997 | 0.5017 | 0.2987 | 0.2788 | 0.2812 |
| 22 | Risk 5 | 20-44 | BMI<30 | Non Smoker | Comorbidity | 0.1055 | 0.0634 | 0.0464 | 0.0476 | 0.4536 | 0.4925 | 0.4944 | 0.2843 | 0.2650 | 0.2673 |
| 4 | Risk 5 | 20-44 | BMI<30 | Non Smoker | No Comorbidity | 0.1401 | 0.0430 | 0.0313 | 0.0321 | 0.4487 | 0.4875 | 0.4894 | 0.2762 | 0.2572 | 0.2595 |
| 3 | Risk 5 | 0-19 | BMI>40 | Non Smoker | No Comorbidity | 0.0008 | 0.0520 | 0.0379 | 0.0389 | 0.2482 | 0.2785 | 0.2800 | 0.1501 | 0.1382 | 0.1396 |
| 2 | Risk 5 | 0-19 | 30<BMI<40 | Non Smoker | No Comorbidity | 0.0010 | 0.0292 | 0.0212 | 0.0217 | 0.2392 | 0.2687 | 0.2703 | 0.1355 | 0.1245 | 0.1258 |
| 1 | Risk 5 | 0-19 | BMI<30 | Non Smoker | No Comorbidity | 0.1463 | 0.0163 | 0.0117 | 0.0120 | 0.2304 | 0.2592 | 0.2607 | 0.1231 | 0.1130 | 0.1142 |

**Table 7.** Profile-stratified $\widehat{P_t(H|I)}$, $\widehat{P_t(Q|H)}$, and $\widehat{P_t(D|Q)}$: Risk profiles (characterized by unique combination of age group, BMI range, smoking status, and any comorbidity), risk group (1-5), model-estimated population prevalence in LAC, the frequency of each profile in the infection population on, and the probability of hospitalization given illness, ICU admission given hospitalization, death given admission to the ICU on May 15, August 1, and October 15, 2020.

with $\mathrm{CFR}(t) > 0.16$; $0.08 < \mathrm{CFR}(t) < 0.16$ in Risk 2; $0.04 < \mathrm{CFR}(t) < 0.08$ in Risk 3; $0.01 < \mathrm{CFR}(t) < 0.04$ in Risk 4; and $\mathrm{CFR}(t) < 0.01$ in Risk 5 (risk groups are indicated for each profile in Table 8) The profile-stratified $\widehat{P_t(H|I)}$, $\widehat{P_t(Q|H)}$, and $\widehat{P_t(D|Q)}$ do not self-order from highest to lowest within the 5 risk groups, since the composite $\mathrm{CFR}(t)$ scales differently than each composing probability for each stage of disease.

### Part III

### Supporting analyses

#### 8 Calculating risk profile stratified CFR and IFR

##### 8.1 CFR/IFR calculation method

We calculate the time-varying risk profile-stratified case fatality rate ($CFR(t)$) and the infection fatality rate ($\mathrm{IFR}(t)$) on the dates May 15, August 1, and October 15, 2020. These estimates are produced from cumulative counts of observed infections, $I_{cum}$, total infections, $I_{cum} + A_{cum}$, and cumulative deaths $D_{cum}$ coming from the epidemic model; and estimates of the time-varying frequency of each risk profile in the infected population, $\mathbf{f}_{t,q,I}$, coming from the risk model 5. The estimated frequency of each profile in the deceased population, $\mathbf{f}_{t,q,D}$, is also required. This is found as the normalized product of the frequency in the incoming stage of disease, the in-ICU population, and the probability of advancing from the ICU to the deceased population, normalized over all risk profiles, i.e.:

$\mathbf{f}_{t,q,D} = \frac{\widehat{\mathbf{Pr(D|Q)}}_{\mathbf{q,t}} \cdot \mathbf{f}_{\mathbf{t,q,Q}}}{\widehat{\mathbf{Pr(D|Q)}}_{\mathbf{q,t}}{}^{\top} \mathbf{f}_{\mathbf{t,q,Q}}}$.

We simulate model realizations with parameter values coming from this joint posterior distribution 2.4. For each realization, we estimate the number of individuals in each risk profile subpopulation in the observed infected population as the estimated frequency of each profile in the infected population, $\mathbf{f}_{t,q,I}$, multiplied by the epidemic-model-estimated time series of the cumulative number of observed infections ($I_{cum}$). Similarly, we estimate the number of individuals in each risk profile subpopulation in the total infected population (including observed an unobserved illnesses) as $\mathbf{f}_{t,q,I}$ multiplied by the estimated time series of cumulative total infections, i.e. $I_{cum} + A_{cum}$. We estimate the number of deceased individuals from each risk profile as $\mathbf{f}_{t,q,D}$ multiplied by the estimated time series of cumulative deaths ($D_{cum}$).

We find the CFR and IFR for each model realization as the estimated number of deaths for each profile over estimated observed infections for each profile, and number of deaths over total infections, respectively. Repeating across the 1000 model realizations achieves the 95% CI.

To produce uncertainty estimates, we aggregate model simulations from 1000 jointly estimated parameter sets. For each parameter set, we aggregate simulations for 500 stochastic epidemic model realizations. We pool together all simulations and report their mean and 2.5th/97.5th percentiles.

##### 8.2 CFR($t$) and IFR($t$) over all risk profiles

The resulting CFR($t$) and IFR($t$) across each risk profile, as well as the estimated frequency of each profile in the overall LAC population and the infected population, each on the dates May 15, August 1, and October 15, 2020, are shown in Table 8.

#### 9 Scenario Analysis

We implement scenarios modifying the population-wide transmission rate at different times and modeling the protection of at-risk populations.

##### 9.1 Modifying the population transmission rate to simulate NPIs

We increase or decrease the value of the reproductive number $R(t)$ to reflect different levels of non-pharmaceutical interventions (NPIs), which could include measures such as physical distancing and/or mask adherence. Specifically, we model three levels of population-wide transmission rates:

1. *NPIs=Observed* implements the observed (epidemic model-estimated) $R(t)$ throughout the epidemic in LAC.
2. *NPIs=Moderate* implements an $R(t)$ equal to the estimated maximum value reached after the initial lockdown restrictions were eased for duration of the epidemic period following the initial decrease. Although this maximum value for $R(t)$ was reached on May 15, in this scenario we decrease $R(t)$ from the initial $R0$ to this maximum $R(t)$ value between March 12 and March 27 to reflect no community lockdown.
3. *NPIs=None* implements $R(t) = R0$ throughout the time interval March - October 2020, representing a baseline scenario in which no actions or behaviors reduce the native $R0$.

The $R(t)$ trend modeled for the three *NPI levels* is shown in Figure 5a in the main text.

##### 9.2 Protecting at-risk populations

To simulate scenarios protecting at-risk populations we focus on the isolation of individuals aged 65+ and calculate values of the probabilities of each stage of disease given infection ($\alpha_t^{Protect.Y}$, $\kappa_t^{Protect.Y}$, and $\delta_t^{Protect.Y}$) over time with $Y\%$ of the subpopulation of individuals 65+ isolated. We isolate a fraction $Y\%$ of individuals 65+ above the observed prevalence of this population in the infected population.

| Profile | Group | age | comorbidity | BMI | smoking | Pop.Prev | Inf.May.15 | Inf.Aug.1 | Inf.Oct.15 | CFR.May.15 | CFR.Aug.1 | CFR.Oct.15 | IFR.May.15 | IFR.Aug.1 | IFR.Oct.15 |
|---|---|---|---|---|---|---|---|---|---|---|---|---|---|---|---|
| 36 | Risk 1 | 65+ | Comorbidity | 30<BMI<40 | Smoker | 0.0002 | 0.0004 | 0.0002 | 0.0002 | 0.2284 | 0.2345 | 0.2250 | 0.0523 | 0.0542 | 0.0521 |
| 30 | Risk 1 | 65+ | Comorbidity | BMI>40 | Non Smoker | 0.0026 | 0.0048 | 0.0027 | 0.0026 | 0.1846 | 0.1907 | 0.1833 | 0.0423 | 0.0441 | 0.0424 |
| 38 | Risk 1 | 45-64 | Comorbidity | BMI>40 | Smoker | 0.0005 | 0.0005 | 0.0005 | 0.0005 | 0.1808 | 0.1862 | 0.1790 | 0.0414 | 0.0430 | 0.0414 |
| 18 | Risk 1 | 65+ | No Comorbidity | 30<BMI<40 | Smoker | 0.0001 | 0.0002 | 0.0001 | 0.0001 | 0.1723 | 0.1718 | 0.1653 | 0.0395 | 0.0397 | 0.0383 |
| 33 | Risk 2 | 65+ | Comorbidity | BMI<30 | Smoker | 0.0019 | 0.0036 | 0.0020 | 0.0019 | 0.1473 | 0.1452 | 0.1398 | 0.0338 | 0.0336 | 0.0324 |
| 12 | Risk 2 | 65+ | No Comorbidity | BMI>40 | Non Smoker | 0.0009 | 0.0016 | 0.0009 | 0.0009 | 0.1389 | 0.1394 | 0.1343 | 0.0318 | 0.0322 | 0.0311 |
| 20 | Risk 2 | 45-64 | No Comorbidity | BMI>40 | Smoker | 0.0003 | 0.0003 | 0.0003 | 0.0003 | 0.1363 | 0.1363 | 0.1313 | 0.0312 | 0.0315 | 0.0304 |
| 27 | Risk 2 | 65+ | Comorbidity | 30<BMI<40 | Non Smoker | 0.0110 | 0.0205 | 0.0114 | 0.0108 | 0.1175 | 0.1164 | 0.1123 | 0.0269 | 0.0269 | 0.0260 |
| 35 | Risk 2 | 45-64 | Comorbidity | 30<BMI<40 | Smoker | 0.0039 | 0.0039 | 0.0042 | 0.0041 | 0.1151 | 0.1136 | 0.1096 | 0.0264 | 0.0263 | 0.0254 |
| 15 | Risk 2 | 65+ | No Comorbidity | BMI<30 | Smoker | 0.0005 | 0.0009 | 0.0005 | 0.0005 | 0.1057 | 0.1020 | 0.0984 | 0.0242 | 0.0236 | 0.0228 |
| 9 | Risk 2 | 65+ | No Comorbidity | 30<BMI<40 | Non Smoker | 0.0037 | 0.0068 | 0.0038 | 0.0036 | 0.0840 | 0.0815 | 0.0787 | 0.0193 | 0.0188 | 0.0182 |
| 17 | Risk 2 | 45-64 | No Comorbidity | 30<BMI<40 | Smoker | 0.0017 | 0.0018 | 0.0019 | 0.0019 | 0.0824 | 0.0796 | 0.0769 | 0.0189 | 0.0184 | 0.0178 |
| 29 | Risk 2 | 45-64 | Comorbidity | BMI>40 | Non Smoker | 0.0090 | 0.0091 | 0.0097 | 0.0096 | 0.0812 | 0.0805 | 0.0779 | 0.0186 | 0.0186 | 0.0180 |
| 37 | Risk 3 | 20-44 | Comorbidity | BMI>40 | Smoker | 0.0013 | 0.0013 | 0.0014 | 0.0013 | 0.0771 | 0.0762 | 0.0737 | 0.0177 | 0.0176 | 0.0170 |
| 24 | Risk 3 | 65+ | Comorbidity | BMI<30 | Non Smoker | 0.0699 | 0.1300 | 0.0722 | 0.0686 | 0.0697 | 0.0670 | 0.0648 | 0.0160 | 0.0155 | 0.0150 |
| 32 | Risk 3 | 45-64 | Comorbidity | BMI<30 | Smoker | 0.0167 | 0.0170 | 0.0180 | 0.0178 | 0.0682 | 0.0653 | 0.0632 | 0.0156 | 0.0151 | 0.0146 |
| 11 | Risk 3 | 45-64 | No Comorbidity | BMI>40 | Non Smoker | 0.0044 | 0.0045 | 0.0048 | 0.0047 | 0.0579 | 0.0561 | 0.0543 | 0.0133 | 0.0130 | 0.0126 |
| 19 | Risk 3 | 20-44 | No Comorbidity | BMI>40 | Smoker | 0.0007 | 0.0007 | 0.0007 | 0.0007 | 0.0550 | 0.0531 | 0.0514 | 0.0126 | 0.0123 | 0.0119 |
| 6 | Risk 3 | 65+ | No Comorbidity | BMI<30 | Non Smoker | 0.0254 | 0.0472 | 0.0262 | 0.0249 | 0.0480 | 0.0455 | 0.0441 | 0.0110 | 0.0105 | 0.0102 |
| 26 | Risk 3 | 45-64 | Comorbidity | 30<BMI<40 | Non Smoker | 0.0382 | 0.0389 | 0.0411 | 0.0408 | 0.0472 | 0.0453 | 0.0439 | 0.0108 | 0.0105 | 0.0102 |
| 14 | Risk 3 | 45-64 | No Comorbidity | BMI<30 | Smoker | 0.0130 | 0.0133 | 0.0140 | 0.0139 | 0.0470 | 0.0443 | 0.0429 | 0.0108 | 0.0102 | 0.0099 |
| 34 | Risk 3 | 20-44 | Comorbidity | 30<BMI<40 | Smoker | 0.0044 | 0.0045 | 0.0048 | 0.0047 | 0.0447 | 0.0427 | 0.0414 | 0.0102 | 0.0099 | 0.0096 |
| 8 | Risk 4 | 45-64 | No Comorbidity | 30<BMI<40 | Non Smoker | 0.0219 | 0.0223 | 0.0236 | 0.0234 | 0.0323 | 0.0306 | 0.0297 | 0.0074 | 0.0071 | 0.0069 |
| 16 | Risk 4 | 20-44 | No Comorbidity | 30<BMI<40 | Smoker | 0.0053 | 0.0054 | 0.0057 | 0.0057 | 0.0306 | 0.0288 | 0.0280 | 0.0070 | 0.0067 | 0.0065 |
| 28 | Risk 4 | 20-44 | Comorbidity | BMI>40 | Non Smoker | 0.0070 | 0.0071 | 0.0075 | 0.0074 | 0.0269 | 0.0258 | 0.0251 | 0.0062 | 0.0060 | 0.0058 |
| 23 | Risk 4 | 45-64 | Comorbidity | BMI<30 | Non Smoker | 0.1510 | 0.1537 | 0.1623 | 0.1609 | 0.0260 | 0.0245 | 0.0238 | 0.0060 | 0.0057 | 0.0055 |
| 31 | Risk 4 | 20-44 | Comorbidity | BMI<30 | Smoker | 0.0206 | 0.0209 | 0.0221 | 0.0219 | 0.0246 | 0.0230 | 0.0224 | 0.0056 | 0.0053 | 0.0052 |
| 10 | Risk 4 | 20-44 | No Comorbidity | BMI>40 | Non Smoker | 0.0073 | 0.0075 | 0.0079 | 0.0078 | 0.0183 | 0.0173 | 0.0169 | 0.0042 | 0.0040 | 0.0039 |
| 5 | Risk 4 | 45-64 | No Comorbidity | BMI<30 | Non Smoker | 0.1045 | 0.1064 | 0.1123 | 0.1114 | 0.0174 | 0.0162 | 0.0157 | 0.0040 | 0.0037 | 0.0036 |
| 13 | Risk 4 | 20-44 | No Comorbidity | BMI<30 | Smoker | 0.0307 | 0.0313 | 0.0330 | 0.0327 | 0.0164 | 0.0152 | 0.0148 | 0.0038 | 0.0035 | 0.0034 |
| 25 | Risk 4 | 20-44 | Comorbidity | 30<BMI<40 | Non Smoker | 0.0238 | 0.0242 | 0.0255 | 0.0253 | 0.0145 | 0.0136 | 0.0132 | 0.0033 | 0.0031 | 0.0031 |
| 7 | Risk 5 | 20-44 | No Comorbidity | 30<BMI<40 | Non Smoker | 0.0240 | 0.0244 | 0.0258 | 0.0255 | 0.0096 | 0.0089 | 0.0087 | 0.0022 | 0.0021 | 0.0020 |
| 22 | Risk 5 | 20-44 | Comorbidity | BMI<30 | Non Smoker | 0.1055 | 0.1074 | 0.1134 | 0.1124 | 0.0075 | 0.0070 | 0.0068 | 0.0017 | 0.0016 | 0.0016 |
| 4 | Risk 5 | 20-44 | No Comorbidity | BMI<30 | Non Smoker | 0.1401 | 0.1427 | 0.1507 | 0.1493 | 0.0049 | 0.0045 | 0.0044 | 0.0011 | 0.0010 | 0.0010 |
| 3 | Risk 5 | 0-19 | No Comorbidity | BMI>40 | Non Smoker | 0.0008 | 0.0002 | 0.0005 | 0.0005 | 0.0018 | 0.0017 | 0.0016 | 0.0004 | 0.0004 | 0.0004 |
| 2 | Risk 5 | 0-19 | No Comorbidity | 30<BMI<40 | Non Smoker | 0.0010 | 0.0002 | 0.0006 | 0.0007 | 0.0009 | 0.0008 | 0.0008 | 0.0002 | 0.0002 | 0.0002 |
| 1 | Risk 5 | 0-19 | No Comorbidity | BMI<30 | Non Smoker | 0.1463 | 0.0346 | 0.0880 | 0.1008 | 0.0004 | 0.0004 | 0.0004 | 0.0001 | 0.0001 | 0.0001 |

**Table 8.** Profile-stratified CFR$(t)$ and IFR$(t)$: Risk profiles (characterized by unique combination of age group, BMI range, smoking status, and any comorbidity), risk group (1-5), model-estimated population prevalence in LAC, the frequency of each profile in the infection population, and the median of the Case Fatality Rate (CFR$(t)$) and Infection Fatality Rate (IFR$(t)$), on May 15, August 1, and October 15, 2020. Profiles with a population prevalence lower than 0.000001 do not yield well-defined CFR or IFR and are not included.

##### *9.2.1 Modeling protection of individuals 65+*

To calculate the probabilities of each stage of disease $\alpha_t^{Protect.Y}$, $\kappa_t^{Protect.Y}$, and $\delta_t^{Protect.Y}$ after isolating (a subgroup of) individuals 65+, we use estimates of the profile-stratified probabilities $\underline{\widehat{P_t(H|I)}}$, $\underline{\widehat{P_t(Q|H)}}$, and $\underline{\widehat{P_t(D|Q)}}$, found in Section 6. We isolate a fraction Y% of individuals 65+ from the estimated frequency of profiles in the infected population, $f_{t,q,I}$, then recursively update the frequency of each profile in each incoming population ($I$, $H$, and $Q$) under the protection scenario. We find the population-average probabilities $\alpha_t^{Protect.Y}$, $\kappa_t^{Protect.Y}$, and $\delta_t^{Protect.Y}$ as the weighted average of the profile-stratified probabilities for each stage of disease from Section 6 and the prevalence of the profiles in the respective incoming population.

Specifically:

We first isolate Y% of individuals 65+ on the two dates determining the different phase of $\alpha_t$, $\kappa_t$, and $\delta_t$, May 1 and August 1, 2020, by removing from $f_{t,q,I}$, the distribution of infections by profile for LAC, Y% of the frequency of all profiles including individuals 65+. After renormalizing to 1, this results in the frequency distribution of profiles in the infected population under the protection scenario $\mathbf{f}_{t,q,I}^{Protect.Y}$.

We can then calculate the population-average probability of advancing from infection to hospitalization under the protection scenario, $\alpha_t^{Protect.Y}$, as

$$\alpha_t^{Protect.Y} = \underline{\widehat{P_t(H|I)}}^{\intercal} \mathbf{f}_{t,q,I}^{Protect.Y}.$$

We find the frequency distribution of profiles in the hospitalized population under the protection scenario $\mathbf{f}_{t,q,H}^{Protect.Y}$ as,

$$\mathbf{f}_{t,q,H}^{Protect.Y} = \frac{\underline{\widehat{P_t(H|I)}} \cdot \mathbf{f}_{t,q,I}^{Protect.Y}}{\underline{\widehat{P_t(H|I)}}^{\mathsf{T}} \mathbf{f}_{t,q,I}^{Protect.Y}}.$$

We then find the population-average probability of advancing from hospitalization to ICU under the protection scenario, $\kappa_t^{Protect.Y}$, as

$$\kappa_t^{Protect.Y} = \underline{\widehat{P_t(Q|H)}}^{\mathsf{T}} \mathbf{f}_{t,q,H}^{Protect.Y}.$$

Finally, we find the frequency distribution of profiles in the in-ICU population under the protection scenario $\mathbf{f}_{t,q,Q}^{Protect.Y}$ as,

$$\mathbf{f}_{t,q,Q}^{Protect.Y} = \frac{\underline{\widehat{P_t(Q|H)}} \cdot \mathbf{f}_{t,q,H}^{Protect.Y}}{\underline{\widehat{P_t(Q|H)}}^{\mathsf{T}} \mathbf{f}_{t,q,H}^{Protect.Y}}.$$

We then find the population-average probability of advancing from hospitalization to ICU under the protection scenario, $\delta_t^{Protect.Y}$, as

$$\delta_t^{Protect.Y} = \underline{\widehat{P_t(D|Q)}}^{\mathsf{T}} \mathbf{f}_{t,q,Q}^{Protect.Y}.$$

###### 9.2.2 Implemented protection scenarios

We model three levels of protection of individuals 65+: (1) *Protect=Observed* models the trend in the probabilities of severe illness estimated by the epidemic model. We call this the "*Protect=Observed*" rather then "*Protect=None*" level because it follows the natural behaviorally-adapted reduction of individuals 65+ in the infected population. (2) *Protect=High* isolates all individuals 65+, approximately 11% of the LAC population [29], from infection and subsequent stages of disease. We implement this scenario by calculating $\alpha_t^{Protect.100}$, $\kappa_t^{Protect.100}$, and $\delta_t^{Protect.100}$ on May 1 and August 1 with individuals 65+ removed. The timeline follows that for the observed trend in $\alpha(t)$, $\kappa(t)$, and $\delta(t)$, with the initial value between March 1 - May 1, a gradual decrease between between May 1 and August 1, and stabilizing at that value for the remainder of the modelled period. (3) *Protect=Moderate* isolates 50% of individuals 65+ from community transmission and subsequent stages of disease. We implement this scenario by calculating $\alpha_t^{Protect.50}$, $\kappa_t^{Protect.50}$, and $\delta_t^{Protect.50}$ on May 1 and August 1 with individuals 65+ removed. The trend of the probabilities of each stage of disease given infection modeled for the three levels is shown in Figure 5b in the main text.

##### 9.3 Simulating scenarios

We implement eight scenarios representing most combinations of the three NPI and three Protect settings. For each scenario, we simulate the model with the estimated parameter values for $R0$, $r$, $T0$, and $p_V$ with the remaining parameters implemented as described above. The model is simulated over the 1000 values of the joint posterior distribution of the estimated parameters with 500 stochastic realizations as described in Section 2.4.

##### 9.4 Scenario results

Figure 3 shows the results of all nine scenarios representing the combinations of the three NPI scenarios, *NPI=Observed, Moderate, Nothing*, and the three protection scenarios, *Protect=Observed, 50%, 100%*. Results for each scenario by CFR(t), IFR(t), cumulative deaths, current in hospital, current observed infections, and current total infections are pictured.

A closer view of Scenarios 4-9 is shown in the main text.

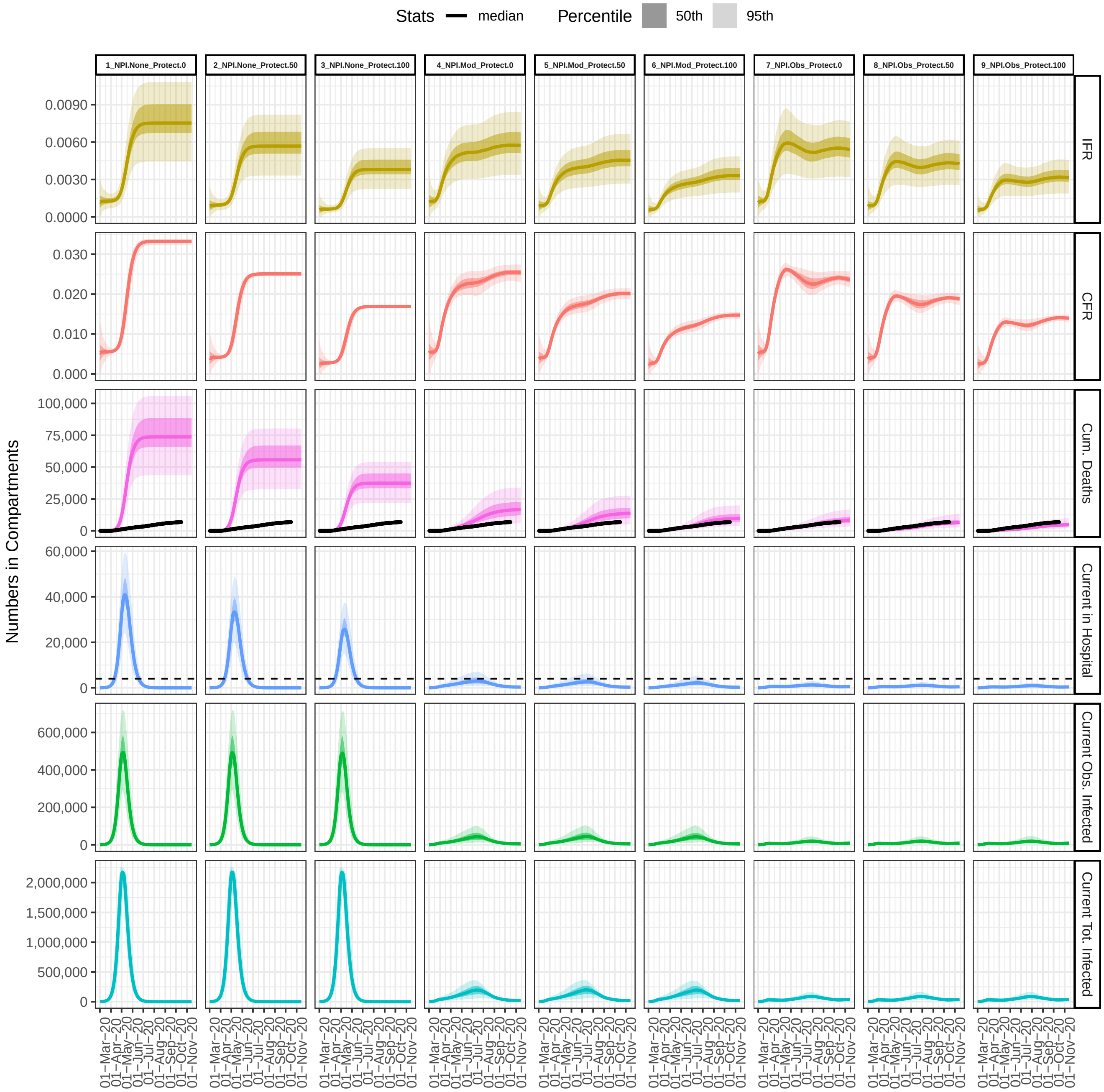

**Figure 3.** The nine scenarios implemented and results by CFR(t), IFR(t), cumulative deaths, current in hospital, current observed infections, and current total infections.
